# Impact of a vaccine passport on first-dose COVID-19 vaccine coverage by age and area-level social determinants in the Canadian provinces of Québec and Ontario: an interrupted time series analysis

**DOI:** 10.1101/2022.10.18.22281192

**Authors:** Jorge Luis Flores Anato, Huiting Ma, Mackenzie A. Hamilton, Yiqing Xia, Sam Harper, David Buckeridge, Marc Brisson, Michael P. Hillmer, Kamil Malikov, Aidin Kerem, Reed Beall, Stefan Baral, Ève Dubé, Sharmistha Mishra, Mathieu Maheu-Giroux

## Abstract

**Background:** In Canada, all provinces implemented vaccine passports in 2021 to increase vaccine uptake and reduce transmission in non-essential indoor spaces. We evaluate the impact of vaccine passport policies on first-dose COVID-19 vaccination coverage by age, area-level income and proportion racialized.

**Methods:** We performed interrupted time-series analyses using vaccine registry data linked to census information in Québec and Ontario (20.5 million people ≥12 years; unit of analysis: dissemination area). We fit negative binomial regressions to weekly first-dose vaccination, using a natural spline to capture pre-announcement trends, adjusting for baseline vaccination coverage (start: July 3^rd^; end: October 23^rd^ Québec, November 13^th^ Ontario). We obtain counterfactual vaccination rates and coverage, and estimated vaccine passports’ impact on vaccination coverage (absolute) and new vaccinations (relative).

**Results:** In both provinces, pre-announcement first-dose vaccination coverage was 82% (≥12 years). The announcement resulted in estimated increases in vaccination coverage of 0.9 percentage points (p.p.;95%CI:0.4-1.2) in Québec and 0.7 p.p. (95%CI:0.5-0.8) in Ontario. In relative terms, these increases correspond to 23% (95%CI:10-36%) and 19% (95%CI:15-22%) more vaccinations. The impact was larger among people aged 12-39 (1-2 p.p.). There was little variability in the absolute impact by area-level income or proportion racialized in either province.

**Conclusions:** In the context of high baseline vaccine coverage across two provinces, the announcement of vaccine passports led to a small impact on first-dose coverage, with little impact on reducing economic and racial inequities in vaccine coverage. Findings suggest the need for other policies to further increase vaccination coverage among lower-income and more racialized neighbourhoods and communities.

**Key messages:**

- Vaccine passport policies increased COVID-19 vaccination coverage by approximately 1 percentage point (19 to 23% increase in vaccinations) in Québec and Ontario, Canada.
- Although vaccine passport policies increased vaccination coverage, absolute gains were limited in the context of high prior vaccine coverage.
- Vaccine passports had little impact on reducing economic and racial inequities in vaccine coverage.

## Introduction

The threat posed by the coronavirus disease (COVID-19) pandemic led to unprecedented interventions, including both generalized and targeted restrictions. Once vaccines became more widely available in 2021 in high-income countries, many governments implemented proof-of-vaccination policies to further mitigate the pandemic’s impacts on population health and the economy.^1^ Often termed “*vaccine passports*”, these policies generally required demonstration of vaccination status or a valid exemption to access non-essential activities and spaces, including restaurants, bars, movie theatres, and concerts.

In Canada, all provinces and the Yukon territory introduced vaccine passports in 2021 and discontinued the policy in the first half of 2022. Québec and Ontario —the two most populous provinces with a population of 22.7 million and Canadian epicenters of the pandemic—^2–4^ were among the first to announce vaccine passports. Provincial governments stated that these policies aimed to reduce SARS-CoV-2 transmission and prevent re-closure of non-essential venues by increasing vaccination coverage and limiting contacts of individuals who had not yet been vaccinated in non-essential venues.^5–7^

The ethical and practical implications of vaccine passports have been debated,^1,8–11^ yet evidence on their effectiveness at incentivizing and increasing coverage of COVID-19 vaccination remains limited. Studies in Europe and Canada found that the introduction of vaccine passports led to increases in vaccination uptake, but this impact depended on age and prior vaccine coverage.^12–14^ These studies have been limited by their use of provincial- or national-level data that restricted exploration of heterogeneity by age and —more importantly— have not examined the effects of vaccine passports according to social determinants of health (SDOH). Given that the COVID-19 pandemic has disproportionately affected communities experiencing social and economic marginalization,^4,15^ it is essential to examine whether vaccination policies resulted in socioeconomic disparities in coverage.

Using vaccine registry data linked to area-level census information, we evaluated the impact of vaccine passports on first-dose vaccination coverage in Québec and Ontario using an interrupted time-series methodology. For each province, we estimated the impact of the vaccine passport by age, and two area-level social determinants: income and proportion racialized.

## Methods

### Study setting and population

In both Québec and Ontario, following early prioritization of residents of long-term care homes and health care workers, vaccination of the general adult population (≥18 years) and youth aged 12-17 years began in May 2021 with BNT162b2 (Pfizer-BioNTech), mRNA-1273 (Moderna), or ChAdOx1 (Oxford-AstraZeneca).^16–19^ COVID-19 proof-of-vaccination policies (herein “vaccine passports”) were announced on August 5^th^ (Québec) and September 1^st^ (Ontario) and came into full force on September 15^th^ (Québec) and September 22^nd^, 2021 (Ontario).^6,7,20^ Non-essential activities and venues targeted by these policies were similar in both provinces (e.g., restaurants, bars, concerts, gyms, school extracurriculars), and restrictions applied to those aged 13 (Québec) or 12 (Ontario) years and over.

### Data sources and measures

We obtained vaccination data from the *Registre de vaccination du Québec* and Ontario’s *COVax* system,^21,22^ which include individuals’ dose administration date, age, and address or dissemination area (DA) of residency. Data were aggregated at the DA level — the smallest standard geographic area for which census information is available (average 400-700 residents).^23^ We included all individuals aged 12 years and over (population eligible for vaccination at time of announcement). Age was categorized based on vaccination priority (12-17, 18-29, 30-39, 40-49, 50-59, and 60+ years). We computed the weekly vaccination rate by DA and age group, defined as the number of first doses administered per 100,000 people without a first dose. We evaluated first-dose coverage because it may better capture people’s response to vaccination mandates, whereas second-dose coverage depended primarily on the time since first dose. Additionally, second-dose eligibility would be affected by changes in the recommended dosage interval (initially 16 weeks but shortened during the summer of 2021).^24–27^

We obtained DA-level after-tax income, per person equivalent from the *Postal Code Conversion File Plus Version 7A/7D*,^28^ and the proportion racialized (based on self-reported “visible minority”) from the latest available Canadian Census (2016) at the time of analysis.^29^ Income was ranked at the census metropolitan area level (to account for within-province variability in cost-of-living) from lowest (1) to highest (5), while proportion racialized was ranked at the provincial level from highest (1) to lowest (5). This ordering was chosen such that the first quintile would align with observed data on the highest incidence of COVID-19 cases.^4,15^ The ranking balanced the population in each quintile (i.e., each quintile had approximately one fifth of the total population).

### Study design and statistical analysis

Analyses were stratified by province. We performed interrupted time-series (ITS) analysis to estimate the impact of the vaccine passport by modeling a counterfactual scenario based on the pre-intervention temporal trend.^30,31^ We allowed for changes in both level and slope of the vaccination rate as a result of the announcement of the vaccine passports. The change lasted for six weeks in both provinces, starting on August 14^th^ (Québec) or September 4^th^ (Ontario). Québec’s date was lagged by one time unit because inspection of the raw data suggested that changes in the weekly rate following the announcement were only detectable after one week, likely because of the announcement timing and decreased vaccination during weekends. The study period was from July 3^rd^ (chosen to align with the end of school year) to five weeks after the end of the vaccine passport’s impact period (i.e., October 23^rd^ for Québec and November 13^th^ for Ontario).

Our modeling approach consisted of two steps. First, we used negative binomial regressions with a natural spline to capture the pre-announcement trend of the DA-level weekly vaccination rates, adjusting for baseline vaccination coverage (i.e., July 3^rd^ 2021; categorical with six groups).^32,33^ Second, we used estimated model coefficients to obtain counterfactual vaccination rates and coverage in the absence of the vaccine passport. We computed the absolute impact of the vaccine passports (observed minus counterfactual vaccination coverage) at the end of the study periods (Québec: October 23^rd^, 2021; Ontario: November 13^th^, 2021). In addition, we calculated the relative increase in vaccine doses administered between the passport announcement and the end of the study period.

We investigated heterogeneity in the impact of vaccine passports by age^12^ and by area-level SDOH, which have been associated with higher COVID-19 infection burden.^4,15^ We fit three models in which the vaccine passport impact could vary by age group, area-level income, or area-level proportion racialized. To further examine trends by SDOH, we fit two additional models with interaction terms between age group and area-level income or proportion racialized. We evaluated heterogeneity of impact by assessing trends in absolute and relative impacts of the vaccine passport by age and SDOH. To examine the impact of vaccine passports on inequities in vaccination coverage, we focused on the magnitude of the absolute impact.

Since heterogeneities in the impact of vaccine passports could be influenced by differences in baseline vaccination coverage, we re-fit the first three models with an interaction term between baseline coverage and the impact of the vaccine passports. We then re-estimated the absolute impact of vaccine passports while holding baseline coverage constant (i.e., setting the baseline variable for all DAs to the same value).

Finally, Montréal and Toronto —the largest census metropolitan areas in each province— were important epicenters of SARS-CoV-2 transmission. Their sociodemographic profiles also differed from the rest of their provinces, which could have affected the impact of vaccine passports. Therefore, main analyses were replicated restricting to the DAs encompassed by these two census metropolitan areas (with the DAs re-ranked according to SDOH).

Confidence intervals (CIs) were obtained using 1,000 bootstrap replicates, using census tracts as the resampling unit to account for geographical and temporal correlations. The 95% CIs were computed by taking the 2.5^th^ and 97.5^th^ quantiles.

### Sensitivity analyses

We explored how alternative modeling choices affected model fit and results by re-parametrizing the age model.^34^ Briefly, we assessed if the chosen start of the study period influenced conclusions by changing the start of the time-series (±1 week), assessed the robustness of our results to different lengths of the vaccine passport impact period (5 or 7 weeks), and examined different model specifications for the time trend (using non-spline methods). Fits were compared based on Akaike Information Criterion, Bayes Information Criterion, and visual assessment.

All analyses were carried out in R V.4.1.0, using packages *fixest* and *splines*.^35–37^ Full details on our modeling approach, model equations, and sensitivity analyses can be found in *Supplementary Materials*.

## Results

### Observed COVID-19 first-dose vaccination coverage over time

In both provinces, first-dose COVID-19 vaccination coverage was 82% in the eligible population (≥12 years) when the vaccine passport was announced. Coverage was highest among people aged 60+ years, with 94% and 87% coverage in Québec and Ontario, respectively, while coverage for those aged 12-17 years was 68% and 76%, respectively. By the end of the study period, vaccination coverage had increased by about 5 percentage points (p.p.) in each province (Table 1).

**Table 1.**
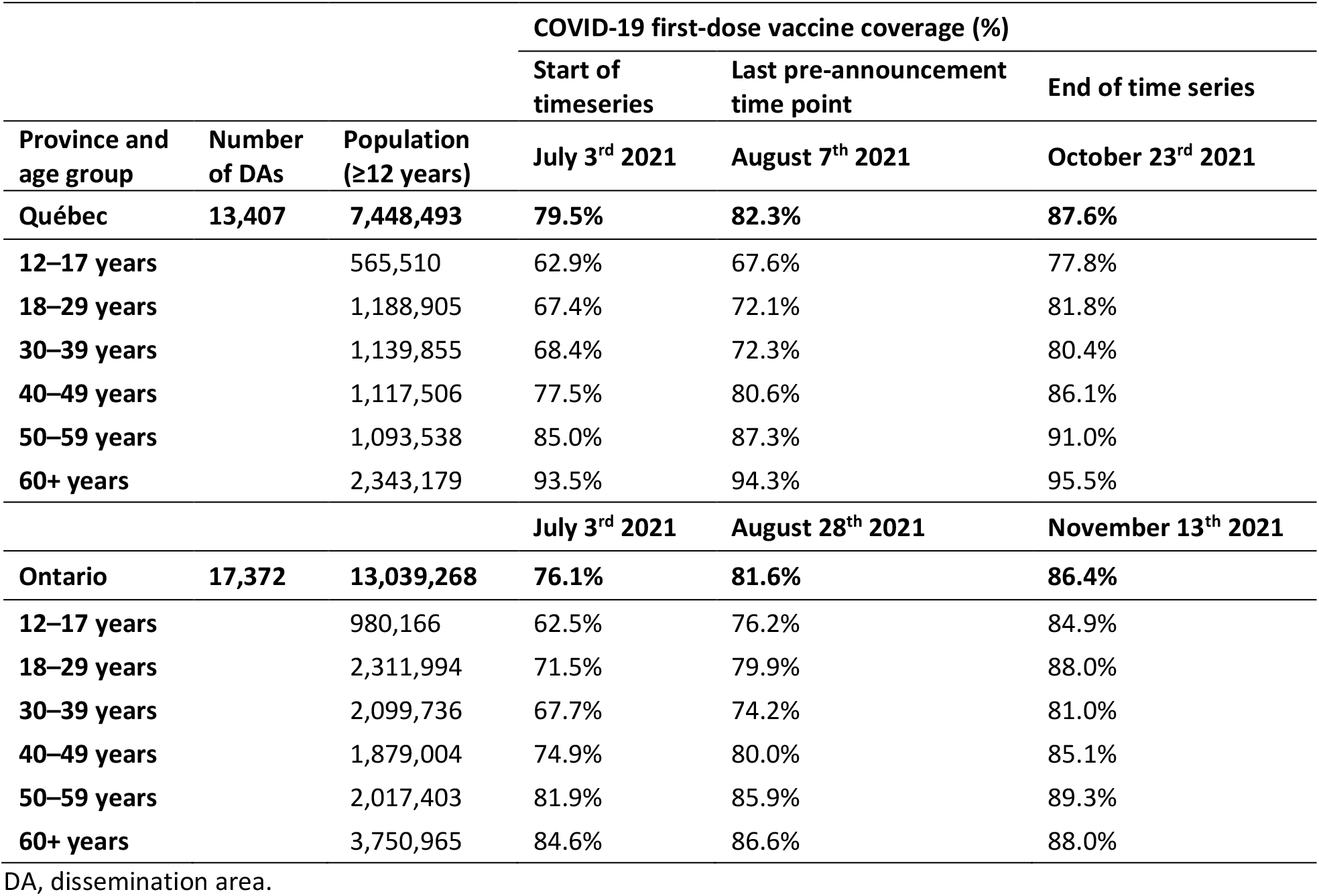
Population sizes and first-dose COVID-19 vaccine coverage for select time points in Québec and Ontario, 2021.

| Province and age group | Number of DAs | Population (≥12 years) | COVID-19 first-dose vaccine coverage (%) |  |  |
| --- | --- | --- | --- | --- | --- |
|  |  |  | Start of timeseries | Last pre-announcement time point | End of time series |
|  |  |  | July 3<sup>rd</sup> 2021 | August 7<sup>th</sup> 2021 | October 23<sup>rd</sup> 2021 |
| **Québec** | **13,407** | **7,448,493** | **79.5%** | **82.3%** | **87.6%** |
| 12–17 years |  | 565,510 | 62.9% | 67.6% | 77.8% |
| 18–29 years |  | 1,188,905 | 67.4% | 72.1% | 81.8% |
| 30–39 years |  | 1,139,855 | 68.4% | 72.3% | 80.4% |
| 40–49 years |  | 1,117,506 | 77.5% | 80.6% | 86.1% |
| 50–59 years |  | 1,093,538 | 85.0% | 87.3% | 91.0% |
| 60+ years |  | 2,343,179 | 93.5% | 94.3% | 95.5% |
|  |  |  | July 3<sup>rd</sup> 2021 | August 28<sup>th</sup> 2021 | November 13<sup>th</sup> 2021 |
| **Ontario** | **17,372** | **13,039,268** | **76.1%** | **81.6%** | **86.4%** |
| 12–17 years |  | 980,166 | 62.5% | 76.2% | 84.9% |
| 18–29 years |  | 2,311,994 | 71.5% | 79.9% | 88.0% |
| 30–39 years |  | 2,099,736 | 67.7% | 74.2% | 81.0% |
| 40–49 years |  | 1,879,004 | 74.9% | 80.0% | 85.1% |
| 50–59 years |  | 2,017,403 | 81.9% | 85.9% | 89.3% |
| 60+ years |  | 3,750,965 | 84.6% | 86.6% | 88.0% |

DA, dissemination area.

Pre-announcement vaccination coverage in the lowest-income DAs was 9 and 7 percentage points (p.p.) lower than highest-income DAs in Québec and Ontario, respectively (similar inequalities in Montréal and Toronto). There were also disparities by proportion racialized: vaccination coverage in DAs with the highest-proportion racialized was 4 p.p. and 8 p.p. lower than the lowest-proportion ones in Québec and Montréal, respectively. In Ontario, these inequalities were reversed — vaccine coverage was 3 p.p. higher in the highest-proportion racialized as compared to the lowest ones, and there was little difference in Toronto (<1p.p.; Supplementary Table S2).

### Observed pre- and post-passport announcement vaccination rates

Prior to the announcement of the vaccine passports, weekly first-dose vaccination rates were stable in Québec and declining in all age groups in Ontario (Figure 1). Increased vaccination rates were observed in both provinces in the week that followed the announcement of the passports, especially among younger age groups (12-17 and 18-29 years old). Comparable increases occurred across income and proportion racialized quintiles. These increases were sustained over a period of six weeks. Similar patterns were observed for Montréal and Toronto (Supplementary Figure S1).

**Figure 1.**
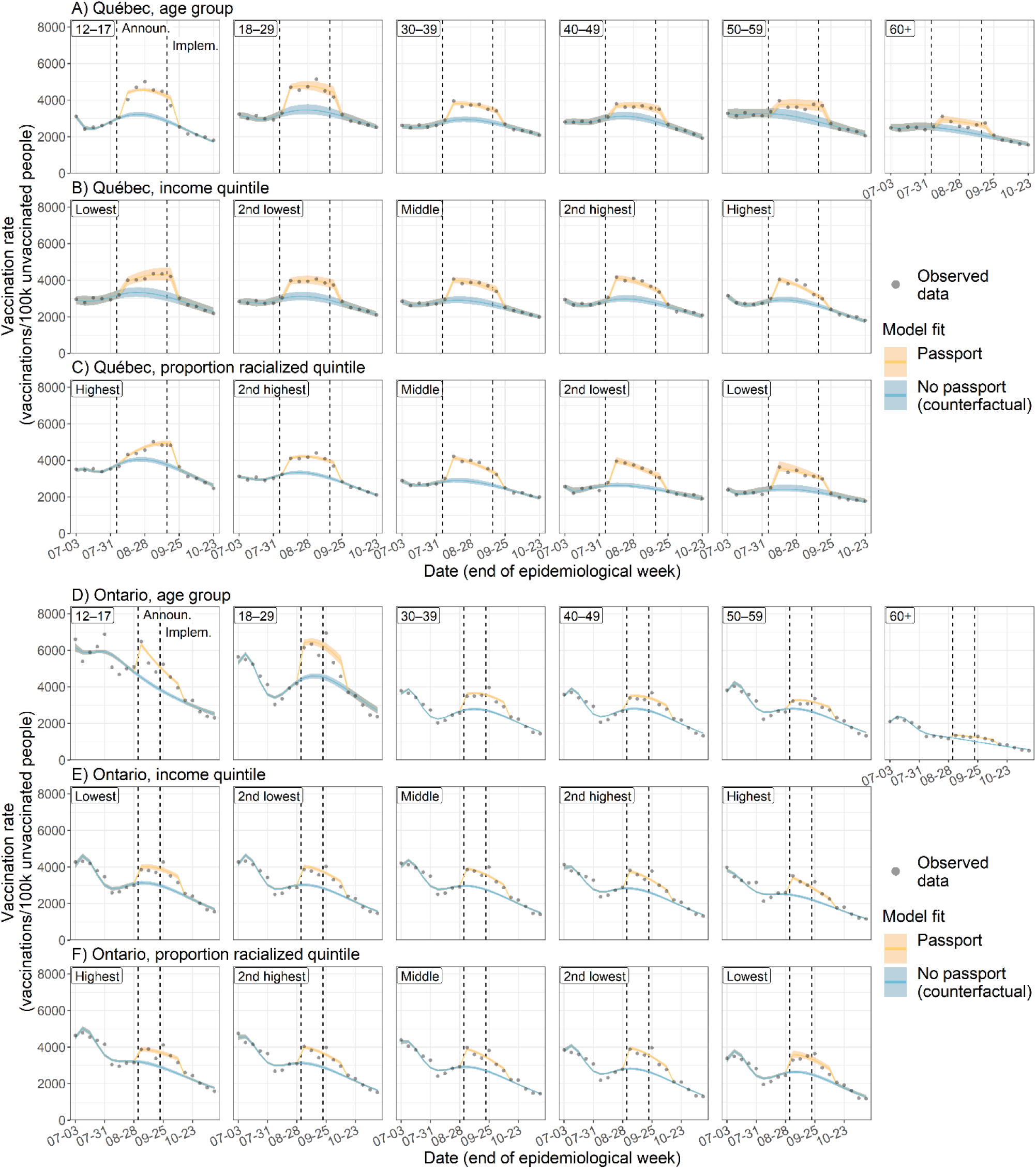
Weekly vaccination rates in Québec (A–C) and Ontario (D–F). Observed (points) and modeled (blue and yellow) vaccination rates over time are shown. Predicted vaccination rates were obtained from three different regression models where the vaccination rate and the impact of the vaccine passport were allowed to vary by age group (A, D), dissemination area (DA)-level income quintile (B, E), or DA-level proportion racialized quintile (C, F). 95% confidence intervals were estimated via bootstrap with 1,000 replicates. Announ., announcement of the vaccine passport; Implem., implementation of the vaccine passport.

### Interrupted time series: vaccine passports’ impact on coverage by age

We estimated that, in the absence of the vaccine passports, first-dose vaccination coverage would have been 0.9 p.p. lower (95%CI: 0.4-1.2) in Québec by October 23^rd^ and 0.7 p.p. lower (95%CI: 0.5-0.8) in Ontario by November 13^th^. In other words, vaccine passports led to relative increases of 23% (Québec; 95%CI: 10-36%) and 19% (Ontario; 95%CI: 15-22%) more vaccinations (Figure 2).

**Figure 2.**
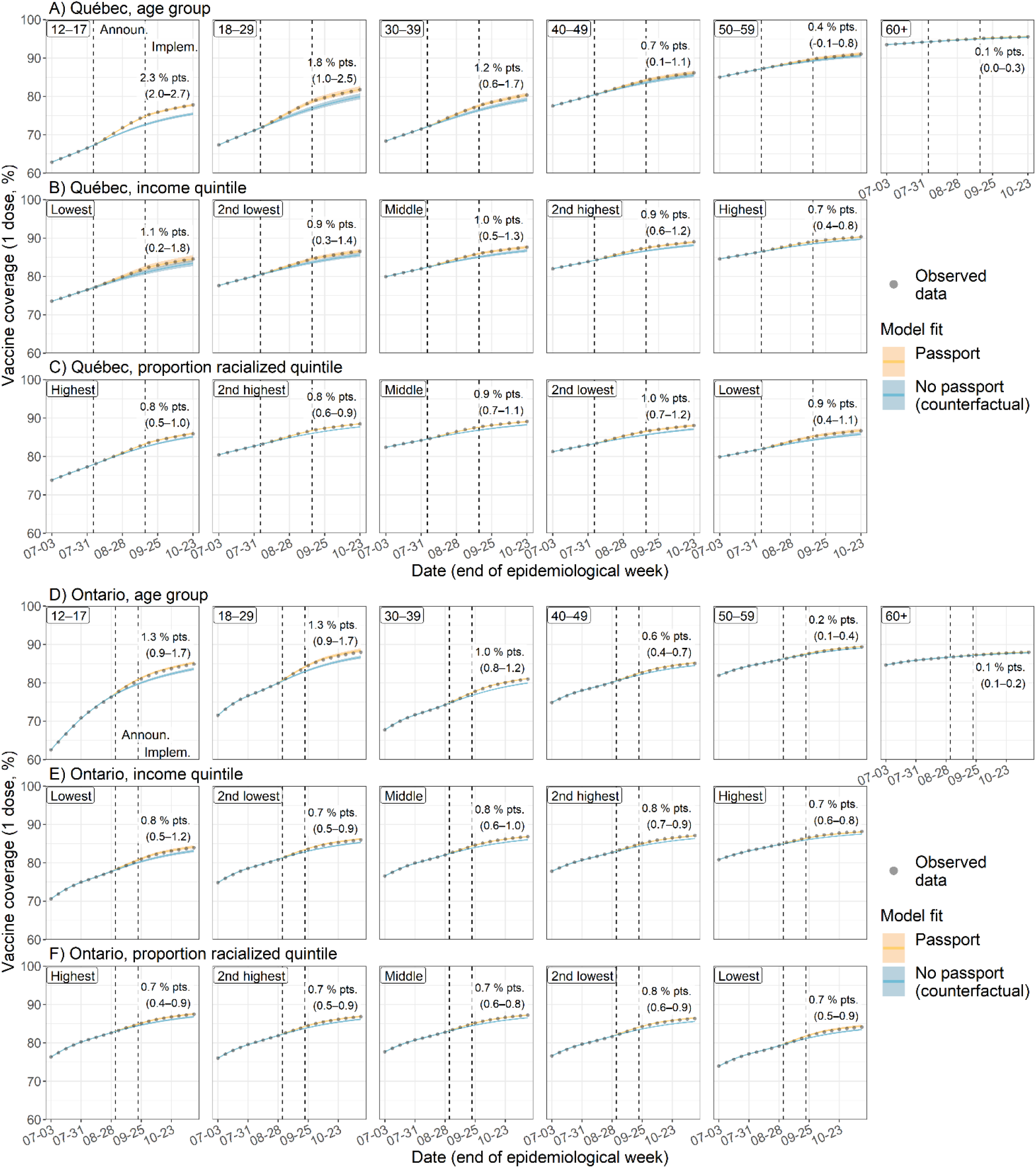
First-dose COVID-19 vaccine coverage in Québec (A–C) and Ontario (D–F). Observed (points) and modeled (blue and yellow) vaccine coverage over time is shown. Predicted vaccine coverage was obtained from three different regression models where the vaccination rate and the impact of the vaccine passport were allowed to vary by age group (A, D), DA-level income quintile (B, E), or DA-level proportion racialized quintile (C, F). Estimates and 95% confidence intervals (CIs) of the impact of the vaccine passport (observed coverage minus modeled counterfactual) are shown at the right of each panel. 95% CIs were estimated via bootstrap with 1,000 replicates. Announ., announcement of the vaccine passport; Implem., implementation of the vaccine passport.

The largest impact of the vaccine passport was observed in the 12-17 age group in Québec, where vaccine coverage was 2.3 p.p. higher (95%CI: 2.0-2.7) than it would have been in the absence of the vaccine passport. In Ontario, the corresponding impact was an increase of 1.3 p.p. (95%CI: 0.9-1.7). The smallest effects were estimated in the 60+ age group, where the impact was around 0.1 p.p. in both provinces (Figure 2A&D). Similar age patterns were observed for the relative impact of the passports (Supplementary Table S2). In Montréal and Toronto, the effect sizes for each age group (except for the 12-17 age group in Toronto) were equivalent to provincial estimates (Supplementary Figure S2). The observed age trends for the absolute impact remained when holding baseline vaccine coverage constant across DAs (Supplementary Figure S3).

### Modification of vaccine passports’ impact on vaccine coverage by age and social determinants of health

When examining the impact by income quintile, we found little evidence of heterogeneity in Québec. In this province, the vaccine passport increased vaccine coverage in the lowest-income DAs by 1.1 p.p. (95%CI: 0.2-1.8) compared to 0.7 p.p. (95%CI: 0.4-0.8) in the highest-income DAs, corresponding to relative increases of 21% (95%CI: 4-40%) and 27% (95%CI: 15-36%), respectively (Figure 2B, Supplementary Table S2). When stratifying by age, the impact of vaccine passports was generally larger in lower-income DAs in most age groups (no clear trend in the 18-29 group). However, uncertainty was large and CIs overlapped across quintiles (Figure 3A).

**Figure 3.**
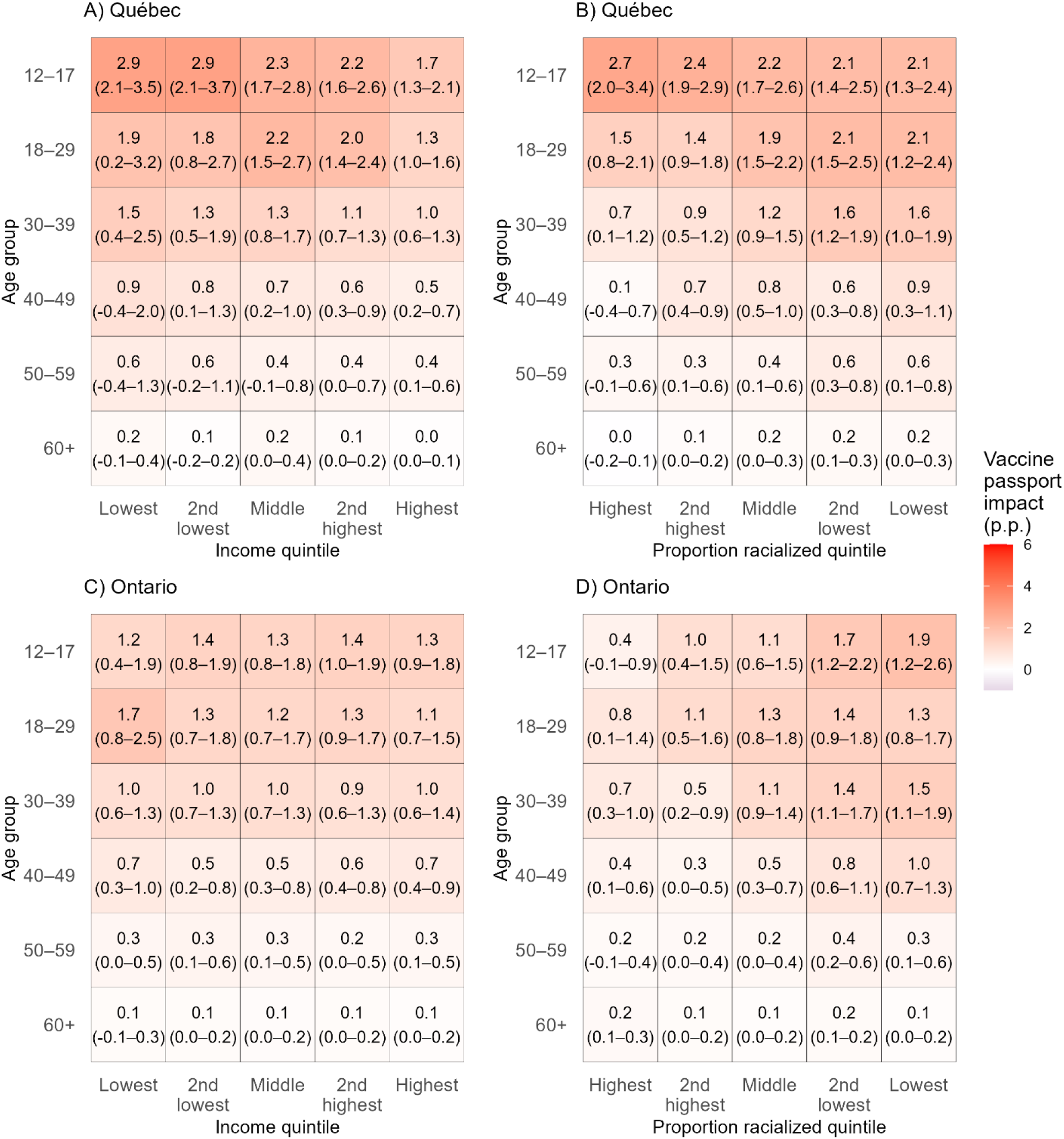
Impact of vaccine passport on first-dose coverage of COVID-19 vaccine (in percentage points) across age and by dissemination area (DA) level of income and proportion of racialized residents in Québec (A, B) and Ontario (C, D) by the end of the study period. The vaccine passport’s impact (defined as the observed vaccination coverage minus the modeled counterfactual coverage in the absence of a vaccine passport) was estimated from two different regression models where the vaccination rate and the impact of the vaccine passport were allowed to vary by the interaction of age and either DA-level income quintile (A, C), or DA-level proportion racialized quintile (B, D). 95% confidence intervals —in parenthesis—were estimated via bootstrap with 1,000 replicates. p.p., percentage points.

The impact was comparable across income quintiles in Ontario at around 0.7 to 0.8 p.p. (Figure 2D), although the relative increase in vaccinations ranged from 19% (95%CI: 10-29%) in the lowest-income DAs to 32% (95%CI: 25-40%) in the highest-income ones (Supplementary Table S2). The lack of heterogeneity in the absolute impact remained with age stratification — the estimated vaccine passport impact was larger in younger age groups but similar across income quintiles within each age group (Figure 3C).

For the proportion racialized, the impact was homogeneous at the DA-level. In Québec, the increase in vaccine coverage was around 0.7 to 1.0 p.p. across quintiles of proportion racialized, with no clear trend (Figure 2C). The relative impact ranged from increases in vaccination of 12% (95%CI: 7.5-18) in DAs with the highest proportion racialized to 29% (95%CI: 10-41%) in the lowest-proportion ones (Supplementary Table S2). Within age groups, the impact was larger in DAs with lower proportion racialized, except for the 12-17 age group where the impact was larger in higher-proportion DAs. Although CIs overlapped across some quintiles, uncertainty was smaller than in the income analyses (Figure 3B).

In Ontario, the DA-level impact was also similar regardless of the proportion of racialized residents. The absolute effect of vaccine passports was 0.7-0.8 p.p. in all quintiles and relative impacts were also homogeneous, ranging from increases of 19% to 24% (Figure 2F, Supplementary Table S2). As in Québec, there was more heterogeneity when stratifying by age, and the impact was bigger in DAs with lower proportion racialized. The effect was attenuated in older age groups, but the gradient remained in all age groups (Figure 3D).

The patterns by income and proportion racialized in Montréal and Toronto were equivalent to those of their respective provinces. One exception was the pattern in the vaccine passport impact by proportion racialized in Toronto, as there was a slight gradient only among people aged 12-29 years (Supplementary Figures S2&S4). When holding baseline vaccination coverage constant, the trends along SDOH remained for all cases except for income in Ontario, where there was a slight gradient in the impact of the vaccine passport (Supplementary Figure S5).

### Sensitivity analyses

In sensitivity analyses (age model), changing the time-series start by ±1 week did not substantially change the estimated effect of the vaccine passport in Québec and slightly lowered it in Ontario (Supplementary Figure S6). In contrast, in models that assumed a different duration for the vaccine passports’ impact (5 or 7 weeks), the impact was slightly lower in both provinces, but model fit was also poorer using these specifications (Supplementary Figures S7-S8). Lastly, when using the best non-spline alternative model specifications (described in Supplementary Methods), the estimated impact of the vaccine passport was slightly higher in Québec and lower in Ontario, as compared to our spline-based approach. All impact estimates were higher when modelled using a simple log-linear model, but these methods had poorer fit (Supplementary Figure S9-S10).

## Discussion

We found that vaccine passports increased COVID-19 first-dose vaccine coverage by approximately 1 p.p. in both Québec and Ontario, where first-dose vaccine coverage was above 80% (≥12-year-old population) at the time passports were announced. This translates to relative increases of 23% (Québec) and 19% (Ontario) more vaccinations among people without a first dose. The impact was largest among younger age groups (<40 years), even after controlling for baseline vaccine coverage. Differences in the impact of vaccine passports by area-level income or proportion of racialized residents were relatively small and the estimates’ uncertainty overlapped, suggesting that vaccine passports had limited impact on reducing socioeconomic disparities in vaccination coverage.

In both provinces, there were inequalities in the pre-announcement vaccination coverage by DA-level income. However, there was only a small gradient in the impact of the vaccine passport in Québec (i.e., higher impact in lower-income DAs), again with overlapping confidence intervals. In Ontario, there was little heterogeneity in the impact of vaccine passports by DA-level income. Taken together, these results suggest that vaccine passports may have led to larger gains in lower-income neighbourhoods in Québec, but not Ontario.

The fact that there were inequalities in baseline vaccine coverage by DA-level proportion of racialized residents in Québec, but not Ontario, could be attributed to different vaccination policies. Québec’s vaccine prioritization focused mostly on age and essential workers, whereas Ontario eventually implemented a “hotspot strategy,” which directed more vaccine-related resources to geographical areas with higher cumulative COVID-19 incidence — which on average had a higher proportion of racialized residents.^38,39^ Although estimates were uncertain, larger absolute effects were observed in neighbourhoods with lower proportions of racialized residents in age-stratified analyses in both provinces. This suggests that vaccine passports may have had slightly larger impacts in predominantly white neighbourhoods despite their higher baseline coverage — a heterogeneity that was masked by differences in age structure and that could manifest in increased disparities in lower-coverage settings.

Our effect size estimates are lower than those previously reported from Europe and Canada.^12–14^ For instance, two studies that evaluated vaccine passports in Italy, France, and Germany found absolute increases in vaccine coverage in the 5-13 p.p. range.^13,14^ In these three countries, however, passports were announced when the fraction of people without a first dose was much larger than Canada (30-35% instead of <20%). In Canada, a study reported slightly higher effects for vaccine passports in Québec (3.1 p.p.) and Ontario (1.9 p.p.).^14^ In contrast to our approach, the authors assumed that vaccine passports would have a permanent effect and positively impact vaccination rates beyond six weeks. Further, they did not account for the continuous reduction in size of the population without first dose, potentially overestimating the impact of vaccine passports.

Our results should also be interpreted by considering vaccine acceptance/hesitancy as a continuum between total acceptance and total refusal.^40,41^ First, vaccine passport policies may have had the biggest impact on those open to vaccination but for whom it was not a priority (i.e., who had scheduled it later in time or were considering it). This could partly explain the observed age effect: younger people may have decided to get vaccinated or moved their vaccinations forward in time to maintain access to non-essential settings and activities targeted by vaccine passports. Second, survey data suggest the majority of residents in Canada expressed positive attitudes toward COVID-19 vaccinations.^42^ Coupled with community-based efforts to improve engagement, increase awareness, and facilitate access (e.g., community ambassador programs, mobile vaccination clinics), there was a large “early adopter” effect by the time the vaccine passports were announced. The remainder of those not yet vaccinated by the time of the vaccine passport announcements may have largely comprised individuals experiencing long-standing, systemic, and persistent barriers to vaccination and/or vaccine mistrust. Our findings suggest that different strategies are needed to address these issues and increase vaccine acceptance and uptake in these communities.^43^

Various limitations should be considered when interpreting these results. First, interrupted time-series methods are sensitive to counterfactual specifications. However, our main results were generally robust to alternative model specifications. Second, concurrent events (e.g., return to school, university-/college-based mandates in Ontario, vaccine lottery in Québec) may have biased estimates of effectiveness upwards. However, school-related events would only partly affect age groups <30, and there is mixed evidence on the impact of vaccine lotteries for COVID-19 vaccination.^44,45^ Third, we used area-level measures of income and racialization, meaning that inferences on the role of individual-level income or racialization could be subject to ecological fallacy. Lastly, this study does not address other ways in which vaccine passports could affect the transmission SARS-CoV-2 (e.g., reduced mixing between people of different vaccination status in non-essential settings). Strengths of our study include the use of detailed DA-level information on vaccinations in Canada’s largest provinces. We also conducted a range of sensitivity analyses that provided credence to our estimates. Lastly, we investigated heterogeneity of impact by age and area-level social determinants of health — known drivers of inequalities in COVID-19 burden.

## Conclusion

In Québec and Ontario, vaccine passports increased COVID-19 vaccination coverage, but absolute gains were small given the provinces already had relatively high vaccination coverage. The impact of vaccine passports was largest among younger age groups in both provinces. However, the effect of vaccine passports varied little by neighbourhood-level SDOH. Ultimately, other policies that account for how social determinants shape barriers to vaccination may be necessary to further increase vaccination coverage and meaningfully reduce inequities in COVID-19-related morbidity and mortality.

## Data Availability

The analysis code is available at https://github.com/pop-health-mod/vaccine-passport-release.
The authors had access to data from the vaccination registries used in this study under agreements with the Institut national de santé publique du Québec and Ontario's Ministry of Health. These data are not available for public release.
The census data used in this study can be downloaded from Statistics Canada at https://www150.statcan.gc.ca/n1/en/catalogue/98-316-X2016001.

## Declarations

### Ethics approval

Ethics approvals were obtained from the *Institutional Review Board* of Faculty of Medicine and Health Sciences of McGill University in Québec (A06-M52-20B) and the *Health Sciences Research Ethics Board* of University of Toronto in Ontario (no. 39253).

### Author contributions

MMG, SM, and JLFA designed the study. JLFA, HM, MAH, MMG, SM, and RB contributed to the conception of the analytical strategy. HM, MAH, and JLFA took care of data management. JLFA carried out all statistical analyses. YX, SH, DB, MB, MPH, KM, AK, RB, SB, and ED all provided substantial input to interpret the results. JLFA drafted the manuscript. All authors contributed to data interpretation and revision of the final manuscript.

### Data availability

The analysis code is available at https://github.com/pop-health-mod/vaccine-passport-release.

The authors had access to data from the vaccination registries used in this study under agreements with the *Institut national de santé publique du Québec* and Ontario’s *Ministry of Health*. These data are not available for public release.

The census data used in this study can be downloaded from Statistics Canada at https://www150.statcan.gc.ca/n1/en/catalogue/98-316-X2016001.

### Supplementary data

Supplementary methods and results are available online.

## Acknowledgements

The authors would like to thank Geneviève Cadieux for her helpful feedback on previous versions of these analyses. MM-G research program is funded by a Canada Research Chair (Tier 2) in Population Health Modeling; and SM research program is funded by a Canada Research Chair (Tier 2) in Mathematical Modeling and Program Science.

## References

1. Katz GM, Born KB, Wit M de, et al. COVID-19 Vaccine Certificates: Key Considerations for the Ontario Context [Internet]. Ontario COVID-19 Science Advisory Table; 2021 Jul. Available from: https://covid19-sciencetable.ca/sciencebrief/covid-19-vaccine-certificates-key-considerations-for-the-ontario-context

2. Xia Y, Ma H, Buckeridge DL, et al. Mortality trends and length of stays among hospitalized patients with COVID-19 in Ontario and Québec (Canada): a population-based cohort study of the first three epidemic waves. Int J Infect Dis. Elsevier; 2022 Aug 1;121:1–10.

3. Godin A, Xia Y, Buckeridge DL, et al. The role of case importation in explaining differences in early SARS-CoV-2 transmission dynamics in Canada—A mathematical modeling study of surveillance data. Int J Infect Dis. Elsevier; 2021 Jan 1;102:254–259.

4. Xia Y, Ma H, Moloney G, et al. Geographic concentration of SARS-CoV-2 cases by social determinants of health in metropolitan areas in Canada: a cross-sectional study. CMAJ. CMAJ; 2022 Feb 14;194(6):E195–E204.

5. Ministère de la Santé et des Services sociaux. Pandémie de la COVID-19 - Un passeport vaccinal pour éviter un confinement généralisé cet automne [Internet]. Salle de presse - MSSS 2021 [cited 2022 Jun 22]. Available from: https://www.msss.gouv.qc.ca/ministere/salle-de-presse/communique-3041/

6. Cabrera H, Olson I. Facing onset of 4th wave of COVID-19 infections, Quebec to implement vaccine passport system. CBC News [Internet]. 2021 Aug 5 [cited 2022 Jun 22]; Available from: https://www.cbc.ca/news/canada/montreal/quebec-vaccine-passport-1.6130699

7. Office of the Premier. Ontario to Require Proof of Vaccination in Select Settings [Internet]. Ontario Newsroom 2021 [cited 2022 Jun 22]. Available from: https://news.ontario.ca/en/release/1000779/ontario-to-require-proof-of-vaccination-in-select-settings

8. Comité d’éthique de santé publique. Avis sur les passeports immunitaires [Internet]. Quebec: INSPQ; 2021 Apr p. 20. Available from: https://www.inspq.qc.ca/publications/3123-avis-passeport-immunitaire-covid19

9. Binks-Collier M. ‘We’re deviating from what makes us Canadian’: An interview with Stefan Baral. Healthy Debate [Internet]. 2021 Nov 25 [cited 2022 Mar 15]; Available from: https://healthydebate.ca/2021/11/topic/stefan-baral-vaccine-passports/

10. Hall MA, Studdert DM. “Vaccine Passport” Certification — Policy and Ethical Considerations. N Engl J Med. 2021 Sep 9;385(11):e32.

11. Bardosh K, Figueiredo A de, Gur-Arie R, et al. The unintended consequences of COVID-19 vaccine policy: why mandates, passports and restrictions may cause more harm than good. BMJ Global Health. BMJ Specialist Journals; 2022 May 1;7(5):e008684.

12. Mills MC, Rüttenauer T. The effect of mandatory COVID-19 certificates on vaccine uptake: synthetic-control modelling of six countries. The Lancet Public Health. Elsevier; 2022 Jan 1;7(1):e15–e22.

13. Oliu-Barton M, Pradelski BSR, Woloszko N, et al. The effect of COVID certificates on vaccine uptake, health outcomes, and the economy. Nat Commun. Nature Publishing Group; 2022 Jul 8;13(1):3942.

14. Karaivanov A, Kim D, Lu SE, Shigeoka H. COVID-19 vaccination mandates and vaccine uptake. Nat Hum Behav. Nature Publishing Group; 2022 Jun 2;1–10.

15. Mishra S, Ma H, Moloney G, et al. Increasing concentration of COVID-19 by socioeconomic determinants and geography in Toronto, Canada: an observational study. Annals of Epidemiology. 2022 Jan 1;65:84–92.

16. Ministère de la Santé et des Services sociaux. Pandémie de la COVID-19 – La vaccination bientôt ouverte à la population générale [Internet]. Salle de presse - MSSS 2021 [cited 2022 Jun 22]. Available from: https://www.msss.gouv.qc.ca/ministere/salle-de-presse/communique-2808/

17. Ministère de la Santé et des Services sociaux. Pandémie de la COVID-19 – La campagne de vaccination des jeunes de 12 à 17 ans s’échelonnera du 25 mai au 23 juin [Internet]. Salle de presse - MSSS 2021 [cited 2022 Jun 22]. Available from: https://www.msss.gouv.qc.ca/ministere/salle-de-presse/communique-2875/

18. Ministry of Health. COVID-19 Vaccine Booking Expanding to Ontarians 18+ Ahead of Schedule [Internet]. Ontario Newsroom 2021 [cited 2022 Jun 22]. Available from: https://news.ontario.ca/en/release/1000143/covid-19-vaccine-booking-expanding-to-ontarians-18-ahead-of-schedule

19. Ministry of Health. COVID-19 Vaccine Booking Expanding to Youth 12+ Ahead of Schedule [Internet]. Ontario Newsroom 2021 [cited 2022 Jun 22]. Available from: https://news.ontario.ca/en/release/1000185/covid-19-vaccine-booking-expanding-to-youth-12-ahead-of-schedule

20. Ministère de la Santé et des Services sociaux. Pandémie de la COVID-19 – Québec dévoile les détails entourant le déploiement du passeport vaccinal [Internet]. Salle de presse - MSSS 2021 [cited 2022 Jun 22]. Available from: https://www.msss.gouv.qc.ca/ministere/salle-de-presse/communique-3112/

21. Registre de vaccination du Québec - À propos [Internet]. Ministère de la Santé et des Services sociaux 2020 [cited 2022 Oct 12]. Available from: https://msss.gouv.qc.ca/professionnels/vaccination/registre-vaccination/

22. Online Access to COVID-19 Vaccination Information for Health Care Providers (HCPs) [Internet]. eHealth Ontario 2021 [cited 2022 Oct 12]. Available from: https://ehealthontario.on.ca/en/news/view/online-access-to-covid-19-vaccination-information-for-hcps

23. Government of Canada SC. Dictionary, Census of Population, 2021 [Internet]. Ottawa, Ontario; 2021 Nov. Report No.: Statistics Canada Catalogue no. 98-301-X. Available from: https://www12.statcan.gc.ca/census-recensement/2021/ref/dict/index-eng.cfm

24. Public Health Agency of Canada. Archived 5: NACI rapid response: Extended dose intervals for COVID-19 vaccines to optimize early vaccine rollout and population protection in Canada [2021-03-03] [Internet]. 2021 [cited 2022 Oct 12]. Available from: https://www.canada.ca/en/public-health/services/immunization/national-advisory-committee-on-immunization-naci/rapid-response-extended-dose-intervals-covid-19-vaccines-early-rollout-population-protection.html

25. Ministère de la Santé et des Services sociaux. Pandémie de la COVID-19 - Un intervalle de 16 semaines entre les deux doses de vaccin - Salle de presse - MSSS [Internet]. Salle de presse - MSSS 2021 [cited 2022 Oct 12]. Available from: https://www.msss.gouv.qc.ca/ministere/salle-de-presse/communique-2676/

26. Ministère de la Santé et des Services sociaux. Pandémie de la COVID-19 – L’intervalle recommandé pour l’ensemble des vaccins administrés au Québec passe de 16 à 8 semaines pour la majorité des personnes - Salle de presse - MSSS [Internet]. Salle de presse - MSSS 2021 [cited 2022 Oct 12]. Available from: https://www.msss.gouv.qc.ca/ministere/salle-de-presse/communique-2916/

27. Office of the Premier. Ontario Accelerates Rollout of Second Shots Targeting a Two-Dose Summer [Internet]. Ontario Newsroom 2021 [cited 2022 Oct 12]. Available from: https://news.ontario.ca/en/release/1000217/ontario-accelerates-rollout-of-second-shots-targeting-a-two-dose-summer

28. Statistics Canada. Postal Code OM Conversion File Plus (PCCF+) [Internet]. [cited 2022 Jul 15]. Available from: https://www150.statcan.gc.ca/n1/en/catalogue/82F0086X

29. Statistics Canada. Census Profile, 2016 [Internet]. 2017. Report No.: Statistics Canada Catalogue no. 98-316-X2016001. Available from: https://www150.statcan.gc.ca/n1/en/catalogue/98-316-X2016001

30. Lopez Bernal J, Cummins S, Gasparrini A. Interrupted time series regression for the evaluation of public health interventions: a tutorial. Int J Epidemiol. 2017 Feb 1;46(1):348–355.

31. Lopez Bernal J, Cummins S, Gasparrini A. Corrigendum to: Interrupted time series regression for the evaluation of public health interventions: a tutorial. Int J Epidemiol. 2021 Jun 1;50(3):1045.

32. Harrell FE. Regression Modeling Strategies: With Applications to Linear Models, Logistic and Ordinal Regression, and Survival Analysis [Internet]. 2nd ed. Springer Cham; 2015 [cited 2022 Jun 22]. Available from: https://link.springer.com/book/10.1007/978-3-319-19425-7

33. Harper S, Bruckner TA. Did the Great Recession increase suicides in the USA? Evidence from an interrupted time-series analysis. Annals of Epidemiology. 2017 Jul 1;27(7):409-414.e6.

34. Lopez Bernal J, Soumerai S, Gasparrini A. A methodological framework for model selection in interrupted time series studies. Journal of Clinical Epidemiology. 2018 Nov 1;103:82–91.

35. R Core Team. R: A language and environment for statistical computing [Internet]. Vienna, Austria: R Foundation for Statistical Computing; 2022 [cited 2022 Jul 18]. Available from: https://www.R-project.org/

36. Bergé L, Krantz S, McDermott G. fixest: Fast Fixed-Effects Estimations [Internet]. 2022 [cited 2022 Aug 9]. Available from: https://CRAN.R-project.org/package=fixest

37. Bergé LR. Efficient estimation of maximum likelihood models with multiple fixed-effects: the R package FENmlm [Internet]. CREA Discussion Papers; 2018 Jul p. 39. Report No.: 13. Available from: http://wwwfr.uni.lu/recherche/fdef/crea/publications2/discussion_papers

38. Mishra S, Stall NM, Ma H, et al. A Vaccination Strategy for Ontario COVID-19 Hotspots and Essential Workers [Internet]. Ontario COVID-19 Science Advisory Table; 2021 Apr. Available from: https://covid19-sciencetable.ca/sciencebrief/a-vaccination-strategy-for-ontario-covid-19-hotspots-and-essential-workers

39. Office of the Premier. Ontario’s COVID-19 Vaccination Strategy Targets High-Risk Neighbourhoods [Internet]. Ontario Newsroom 2021 [cited 2022 Aug 3]. Available from: https://news.ontario.ca/en/release/61124/ontarios-covid-19-vaccination-strategy-targets-high-risk-neighbourhoods

40. MacDonald NE, Comeau J, Dubé È, et al. Royal society of Canada COVID-19 report: Enhancing COVID-19 vaccine acceptance in Canada. FACETS. Canadian Science Publishing; 2021 Jan;6:1184–1246.

41. Dubé È, Ward JK, Verger P, MacDonald NE. Vaccine Hesitancy, Acceptance, and Anti-Vaccination: Trends and Future Prospects for Public Health. Annu Rev Public Health. 2021 Apr 1;42(1):175–191.

42. Dionne M, Dubé È, Hamel D, Rochette L, Tessier M. Pandémie et vaccination contre la COVID-19 - 9 août 2022 [Internet]. Institut national de santé publique du Québec 2022 [cited 2022 Sep 19]. Available from: https://www.inspq.qc.ca/covid-19/sondages-attitudes-comportements-quebecois/vaccination-09-aout-2022

43. Dubé E, Gagnon D, MacDonald NE, the SAGE Working Group on Vaccine Hesitancy. Strategies intended to address vaccine hesitancy: Review of published reviews. Vaccine. 2015 Aug 14;33(34):4191–4203.

44. Dave D, Friedson AI, Hansen B, Sabia JJ. Association Between Statewide COVID-19 Lottery Announcements and Vaccinations. JAMA Health Forum. 2021 Oct 15;2(10):e213117.

45. Acharya B, Dhakal C. Implementation of State Vaccine Incentive Lottery Programs and Uptake of COVID-19 Vaccinations in the United States. JAMA Network Open. 2021 Dec 9;4(12):e2138238.

