## Supplementary Materials for "Impact of a vaccine passport on first-dose COVID-19 vaccine coverage by age and area-level social determinants in the Canadian provinces of Québec and Ontario: an interrupted time series analysis"

### Supplementary methods

#### Data sources and measures

| Variables | Source | Definition |
|---|---|---|
| **Population** | Registered-persons databases | Number of people of a given age group in a dissemination area (DA), excluding long-term care home residents. These numbers were adjusted for over-vaccination through the following heuristic: if there were more doses than people in a given age group and DA by the end of the timeseries (Québec: October 23rd 2021; Ontario: November 13th 2021), we set the population size to the number of doses observed on that date. |
| **After-tax income, per person equivalent (100% of census sample)** | Postal Code Conversion File Plus Version 7A/7D | After-tax income is calculated for each household from the income for all household members. Calendar year 2015 is the reference period for all income variables in the 2016 Census. Single-person equivalent is used to account for households of different sizes. To account for differences in the cost of living, the ranking is calculated exclusively from DAs within the same census metropolitan area. |
| **Proportion of visible minority (proportion racialized in the main text)** | 2016 Canadian Census *(25% of census sample)* | Visible minority refers to a person's self-identification as a visible minority as defined by the *Employment Equity Act*: "persons, other than Aboriginal peoples, who are non-Caucasian in race or non-white in colour." According to the 2021 Census Dictionary, "the visible minority population consists mainly of the following groups: South Asian, Chinese, Black, Filipino, Arab, Latin American, Southeast Asian, West Asian, Korean and Japanese." |

#### Identification strategy: interrupted time series

We use an interrupted time series approach to identify the impact of vaccine passports on vaccine coverage. This was preferred to a difference-in-difference analysis because of provincial differences in trends in vaccination rates prior to the announcement of vaccine passports (i.e., stable in Québec and decreasing in Ontario, violating the parallel trends assumption). For each province, we modeled the dissemination area (DA)-level weekly vaccination rate using negative binomial regression models to account for overdispersion (Supplementary Figures S11-S13), with an offset term for the population size without a first dose. Calendar time was modeled using a natural spline (i.e., restricted cubic spline), with three knots placed at the 10th, 50th and 90th quantiles of the pre-announcement period.[1,2] Because pre-announcement vaccine coverage could influence subsequent weekly vaccination rates, we adjusted for DA-level vaccine coverage (age-stratified, except for models 2a and 2b which used vaccine data for the whole DA population) at the start of the timeseries (i.e., July 3rd 2021). Vaccine coverage at the start of the timeseries (July 3rd) was categorized into six groups: <50%, 50%–<60%, 60%–<70%,70%–<80%, 80–<90%, and ≥90%.

For the interrupted time series, the impact of the vaccine passport was modeled to have an immediate change in the level and slope of the vaccination rate—as measured by regression coefficients $\beta$ and $\delta$ in the regression formulas (see *Equations for the interrupted time series analyses*). Based on inspection of the raw data, vaccine passports had transitory effects on vaccination rates. The length of this impact period was determined empirically via model comparison (Akaike Information Criterion, Bayes Information Criterion, and visual assessment). In both provinces, the best fit was provided by an impact period of 6 weeks, starting on August 14th (Québec) or September 4th (Ontario).

**Equations for the interrupted time series analyses**

*Main model: weekly first-dose vaccination rates ($\boldsymbol{\lambda_{i,a,t}}$) in DA i, for age group* a*, at time* t *are modeled on the logarithmic scale.*

The main age-stratified model (model 1) takes the following form

$$\log(\lambda_{i,a,t}) = \alpha + f(T_t) + \beta * P_t + \delta * (T_t - I) * P_t + \\ \sum[\gamma_a + f(T_t) + \beta_a * P_t + \delta_a * (T_t - I) * P_t] * A_a + \sum(\psi_a * C_{i,a}) + offset(p_{i,a,t})$$

Where $\alpha$ is the model intercept; $f(T_t)$ is the natural cubic spline function for calendar time in weeks ($T_t$); $\beta$ is the level change (for the reference group) in log vaccination rate during the time period ($P_t$) over which the vaccine passports is presumed to have an effect; $\delta$ is the coefficient for the slope change (for the reference group) in log vaccination rates when the vaccine passports have an effect and $I$ indicates the week during which vaccine passports are announced; $\gamma_a$ is a vector of coefficients corresponding to age group a and $A_a$ is a vector containing the dummy variables for age; similarly, $\beta_a$ is a vector containing the age-specific coefficients for the level change in vaccination rates and $\delta_a$ the vector of coefficients for the age-specific slope changes; finally, $\psi_a$ contains the coefficients for differences in baseline vaccine coverage on vaccination rates; $C_{i,a}$ is a matrix of containing the dummy variables for vaccine coverage in DA *i* and age group *a* and $p_{i,a,t}$ is the population size without a first dose in DA *i*, for age group *a* and at time *t*.

*Models with dissemination area-level income or proportion racialized*

The equation above can be adapted to examine if vaccine passports had a differential impact by income quintile (model 2a). In this case, the $s$ subscript indicates the income quintile of DA *i* and $S_i$ is a vector that contains the dummy variables for the income quintiles. The equation is the following:

$$\log(\lambda_{i,t}) = \alpha + f(T_t) + \beta * P_t + \delta * (T_t - I) * P_t + \sum[\gamma_s + f(T_t) + \beta_s * P_t + \delta_s * (T_t - I) * P_t] * S_i + \sum(\psi * C_i) + offset(p_{i,t})$$

Similarly, for the DA-level proportion of racialized residents, the equation above is replaced with the one below (model 2b). The differences being that the *v* indicate the proportion racialized quintiles and $V_i$ is a vector of the dummy variables for proportion racialized quintiles.

$$\log(\lambda_{i,t}) = \alpha + f(T_t) + \beta * P_t + \delta * (T_t - I) * P_t + \sum[\gamma_v + f(T_t) + \beta_v * P_t + \delta_v * (T_t - I) * P_t] * V_i + \sum(\psi * C_i) + offset(p_{i,t})$$

*Models with interaction terms between age and dissemination area-level income or proportion of racialized residents*

The interrupted time series models above can be modified to examine if vaccine passports have differential impact by age and our two social determinants of health (i.e., interaction). The notation above applies here and the equation for the model with interactions between age and income quintiles (model 3a) is the following:

$$\begin{aligned}\log(\lambda_{i,a,t}) = {} & \alpha + f(T_t) + \beta * P_t + \delta * (T_t - I) * P_t + \sum(\psi_a * C_{i,a}) + \\ & \sum[\gamma_a + f(T_t) + \beta_a * P_t + \delta_a * (T_t - I) * P_t] * A_a + \\ & \sum[\gamma_s + f(T_t) + \beta_s * P_t + \delta_s * (T_t - I) * P_t] * S_i + \\ & \sum[\gamma_{a,s} + f(T_t) + \beta_{a,s} * P_t + \delta_{a,s} * (T_t - I) * P_t] * A_a * S_i + offset(p_{i,a,t})\end{aligned}$$

Finally, the equation for the model with interactions between age and quintiles of the proportion racialized (model 3b) is:

$$\begin{aligned}\log(\lambda_{i,a,t}) = {} & \alpha + f(T_t) + \beta * P_t + \delta * (T_t - I) * P_t + \sum(\psi_a * C_{i,a}) + \\ & \sum[\gamma_a + f(T_t) + \beta_a * P_t + \delta_a * (T_t - I) * P_t] * A_a + \\ & \sum[\gamma_v + f(T_t) + \beta_v * P_t + \delta_v * (T_t - I) * P_t] * V_i + \\ & \sum[\gamma_{a,v} + f(T_t) + \beta_{a,v} * P_t + \delta_{a,v} * (T_t - I) * P_t] * A_a * V_i + offset(p_{i,a,t})\end{aligned}$$

In these last two equations, the *a,s* and *a,v* subscripts indicate that the variables reference matrices of coefficients (instead of vectors) in which rows correspond to the income or proportion racialized quintile, respectively, and columns correspond to the age group.

**Sensitivity analyses**

Inferences from interrupted time series can be sensitive to the modeling of the counterfactual scenario.[3] We performed three different sensitivity analyses to estimate how alternative modeling choices would affect model fit, results, and conclusions.

First, we determined whether the estimated counterfactual (and thus, estimated impact of the vaccine passport) was sensitive to changing the starting date of the time series. We compared the starting date of July 3rd to models starting one week earlier (June 26th) or one week later (July 10th).

Second, we determined whether the counterfactual was sensitive to changing the length of the impact period – i.e., the length during which the $P_t$ variable equals 1– of the vaccine passport. We compared the best-fitting model of six weeks to models with an impact period of either five or seven weeks.

Lastly, we tested whether simpler model specifications for calendar time would be able to capture the observed vaccination rates in Québec and Ontario and replicate our spline-based results. Given the different trends observed in vaccination rates over the summer, we used different models for each province (which we determined heuristically to fit the data well). For Québec, we modeled the log rate-time relationship with a linear trend and an indicator variable for the time period after the impact of the vaccine passport, i.e., after September 18th. In Ontario, we modeled the relationship with linear and quadratic terms for time, and an indicator variable for the month of July. We also tested a simple model in which the relationship between log vaccination rate and time is modeled via a simple linear relationship, to test whether this simpler model appropriately captured the pre-announcement vaccination trend in each province. The equations for these alternative models are provided below.

*Best alternative model specifications*

For both provinces, the best non-spline alternative model specifications are the same as the age-stratified model 1, except for how the time trend is modeled.

For Québec, we replace the spline for time $f(T_t)$ by a coefficient for time (in weeks) $\eta$ and $\eta_a$. We also allow the intercept and slope of the log vaccination rate after the vaccine passport impact “wears off” to differ from the pre-announcement vaccination rate, via coefficients $\theta$, $\theta_a$, $\kappa$ and $\kappa_a$. The time period after which the vaccine passport is presumed to have an effect is denoted by $R_t$. As before, coefficients without a subscript are the coefficient for the reference age group, and coefficients with the subscript *a* are a vector of coefficients for the remaining age groups.

$$\log(\lambda_{i,a,t}) = \alpha + \eta * T_t + \beta * P_t + \delta * (T_t - I) * P_t + \theta * R_t + \kappa * (T_t - I - 6) * R_t + \sum[\theta_a * R_t + \kappa_a * (T_t - I - 6) * R_t] * A_a + \sum[\gamma_a + \eta_a * T_t + \beta_a * P_t + \delta_a * (T_t - I) * P_t] * A_a + \sum(\psi_a * C_{i,a}) + offset(p_{i,a,t})$$

For Ontario, the coefficients $\theta$, $\theta_a$, $\kappa$ and $\kappa_a$ are used to allow the intercept and slope of the vaccination rate to differ in July, denoted by the indicator variable $J_t$. The coefficients for the quadratic term are $\omega$ and $\omega_a$.

$$\log(\lambda_{i,a,t}) = \alpha + \eta * T_t + \beta * P_t + \delta * (T_t - I) * P_t + \theta * J_t + \kappa * T_t * J_t + \omega * T_t + \sum[\theta_a * J_t + \kappa_a * T_t * J_t + \omega_a * T_t] * A_a + \sum[\gamma_a + \eta_a * T_t + \beta_a * P_t + \delta_a * (T_t - I) * P_t] * A_a + \sum(\psi_a * C_{i,a}) + offset(p_{i,a,t})$$

*Linear model between log vaccination rate and time*

The linear models are as above, except that the spline for time is only replaced by the $\eta$ and $\eta_a$ coefficients. No other coefficients or quadratic terms are used.

$$\log(\lambda_{i,a,t}) = \alpha + \eta * T_t + \beta * P_t + \delta * (T_t - I) * P_t + \sum[\gamma_a + \eta_a * T_t + \beta_a * P_t + \delta_a * (T_t - I) * P_t] * A_a$$

**References for the supplementary methods**

1. Harrell FE. Regression Modeling Strategies: With Applications to Linear Models, Logistic and Ordinal Regression, and Survival Analysis [Internet]. 2nd ed. Springer Cham; 2015 [cited 2022 Jun 22]. Available from: https://link.springer.com/book/10.1007/978-3-319-19425-7

2. Harper S, Bruckner TA. Did the Great Recession increase suicides in the USA? Evidence from an interrupted time-series analysis. *Annals of Epidemiology*. 2017 Jul 1;**27**(7):409-414.e6.

3. Lopez Bernal J, Soumerai S, Gasparrini A. A methodological framework for model selection in interrupted time series studies. *Journal of Clinical Epidemiology*. 2018 Nov 1;**103**:82–91.

### Supplementary results

**Supplementary Table S1. Population sizes and first-dose COVID-19 vaccine coverage for select time points in the Montréal and Toronto census metropolitan areas, 2021.**

| | | | COVID-19 first-dose vaccine coverage (%) | | |
|---|---|---|---|---|---|
| | | | Start of timeseries | Last pre-announcement time point | End of time series |
| **Province and age group** | **Number of DAs** | **Population (≥12 years)** | **July 3rd 2021** | **August 7th 2021** | **October 23rd 2021** |
| **Québec** | **6,437** | **3,696,253** | **78.9%** | **82.2%** | **88.1%** |
| **12–17 years** | | 299,847 | 60.0% | 65.2% | 76.3% |
| **18–29 years** | | 639,506 | 69.1% | 74.1% | 84.1% |
| **30–39 years** | | 600,349 | 69.5% | 73.9% | 82.2% |
| **40–49 years** | | 584,213 | 78.3% | 81.8% | 87.8% |
| **50–59 years** | | 543,385 | 85.2% | 87.9% | 91.9% |
| **60+ years** | | 1,028,953 | 93.2% | 94.1% | 95.6% |
| | | | **July 3rd 2021** | **August 28th 2021** | **November 13th 2021** |
| **Ontario** | **7,333** | **5,777,554** | **77.4%** | **82.8%** | **87.2%** |
| **12–17 years** | | 442,406 | 65.2% | 78.9% | 86.6% |
| **18–29 years** | | 1,080,806 | 77.1% | 84.8% | 91.9% |
| **30–39 years** | | 991,693 | 72.1% | 78.1% | 83.8% |
| **40–49 years** | | 881,175 | 77.0% | 81.7% | 86.1% |
| **50–59 years** | | 903,509 | 81.9% | 85.7% | 88.9% |
| **60+ years** | | 1,477,965 | 82.2% | 84.4% | 86.0% |

DA, dissemination area.

**Supplementary Table S2. First-dose COVID-19 vaccination coverage before the vaccine passport announcement, absolute and relative impact of the vaccine passport in Québec and Ontario by age, income quintile, and proportion racialized quintile.**

| | **Québec** | | |
|---|---|---|---|
| | **Pre-announcement vaccine coverage (%)** | **Absolute impact (p.p. change in coverage)** | **Relative impact (% increase in doses)** |
| **Age** | | | |
| 12–17 | 67.6 | 2.3 (2.0–2.7) | 36 (28–43) |
| 18–29 | 72.1 | 1.8 (1.0–2.5) | 28 (13–43) |
| 30–39 | 72.3 | 1.2 (0.6–1.7) | 22 (9.9–33) |
| 40–49 | 80.6 | 0.7 (0.1–1.1) | 16 (2.3–30) |
| 50–59 | 87.3 | 0.4 (-0.1–0.8) | 16 (-1.7–36) |
| 60+ | 94.3 | 0.1 (0.0–0.3) | 14 (-2.7–32) |
| **Income quintile** | | | |
| *Lowest* | 77.3 | 1.1 (0.2–1.8) | 21 (3.8–40) |
| *2nd lowest* | 80.6 | 0.9 (0.3–1.4) | 22 (6.8–38) |
| *Middle* | 82.5 | 1.0 (0.5–1.3) | 28 (13–42) |
| *2nd highest* | 84.4 | 0.9 (0.6–1.2) | 30 (16–42) |
| *Highest* | 86.6 | 0.7 (0.4–0.8) | 27 (15–36) |
| **Proportion racialized quintile** | | | |
| *Highest* | 78.1 | 0.8 (0.5–1.0) | 12 (7.5–18) |
| *2nd highest* | 83.2 | 0.8 (0.6–0.9) | 21 (16–26) |
| *Middle* | 84.7 | 0.9 (0.7–1.1) | 30 (22–39) |
| *2nd lowest* | 83.5 | 1.0 (0.7–1.2) | 33 (20–42) |
| *Lowest* | 82.0 | 0.9 (0.4–1.1) | 29 (9.8–41) |

| | **Ontario** | | |
|---|---|---|---|
| | **Pre-announcement vaccine coverage (%)** | **Absolute impact (p.p. change in coverage)** | **Relative impact (% increase in doses)** |
| **Age** | | | |
| 12–17 | 76.2 | 1.3 (0.9–1.7) | 22 (14–30) |
| 18–29 | 79.9 | 1.3 (0.9–1.7) | 24 (15–33) |
| 30–39 | 74.2 | 1.0 (0.8–1.2) | 21 (17–25) |
| 40–49 | 80.0 | 0.6 (0.4–0.7) | 15 (11–20) |
| 50–59 | 85.9 | 0.2 (0.1–0.4) | 9.2 (4.8–14) |
| 60+ | 86.6 | 0.1 (0.1–0.2) | 12 (6.2–18) |
| **Income quintile** | | | |
| *Lowest* | 77.7 | 0.8 (0.5–1.2) | 19 (9.5–29) |
| *2nd lowest* | 80.8 | 0.7 (0.5–0.9) | 18 (12–25) |
| *Middle* | 82.0 | 0.8 (0.6–1.0) | 24 (18–31) |
| *2nd highest* | 82.7 | 0.8 (0.7–0.9) | 27 (21–33) |
| *Highest* | 84.8 | 0.7 (0.6–0.8) | 32 (25–40) |
| **Proportion racialized quintile** | | | |
| *Highest* | 82.6 | 0.7 (0.4–0.9) | 19 (11–27) |
| *2nd highest* | 81.9 | 0.7 (0.5–0.9) | 19 (13–26) |
| *Middle* | 82.8 | 0.7 (0.6–0.8) | 23 (17–29) |
| *2nd lowest* | 81.6 | 0.8 (0.6–0.9) | 24 (19–29) |
| *Lowest* | 79.1 | 0.7 (0.5–0.9) | 19 (12–25) |

The pre-announcement vaccination coverage corresponds to the coverage as of August 7th (Québec) or August 28th (Ontario), 2021. Absolute impact was estimated as observed coverage minus modeled counterfactual by the end of the study period, October 23rd (Québec) or November 13th (Ontario), 2021. The relative impact was estimated as the observed number of doses administered divided by the modeled counterfactual number of doses, during the period between the passport announcement and the end of the study period. Columns for the estimated impact show the point estimate and the 95% confidence intervals (CIs) in parenthesis. 95% CIs were estimated via bootstrap with 1,000 replicates.

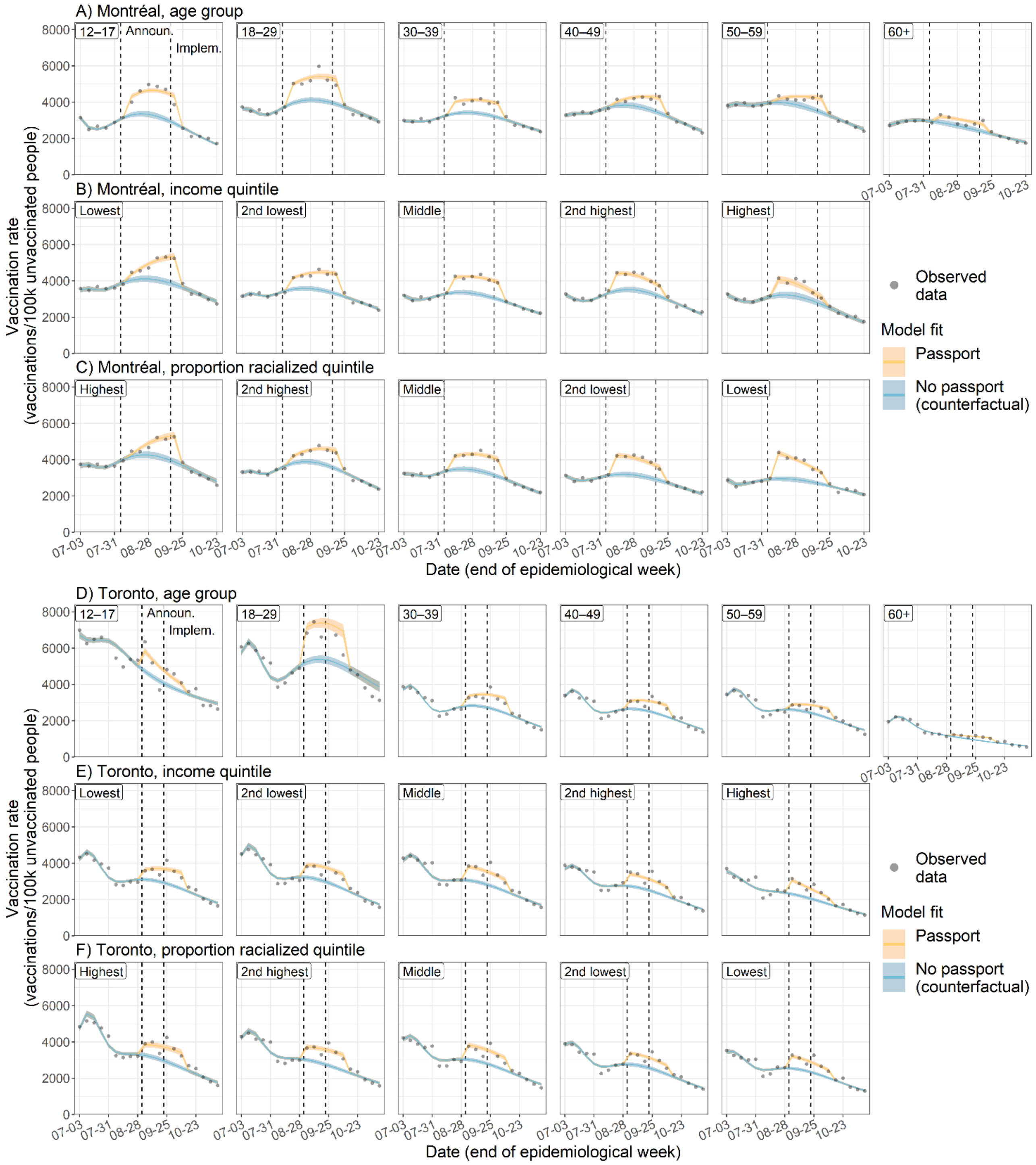


**Supplementary Figure S1. Weekly vaccination rates in Montréal (A–C) and Toronto (D–F).** Observed (points) and modeled (blue and yellow) vaccination rates over time are shown. Predicted vaccination rates were obtained from three different regression models where the vaccination rate and the impact of the vaccine passport were allowed to vary by age group (A, D), DA-level income quintile (B, E), or DA-level proportion racialized quintile (C, F). 95% confidence intervals were estimated via bootstrap with 1,000 replicates. (Announ., announcement of the vaccine passport; Implem., implementation of the vaccine passport)

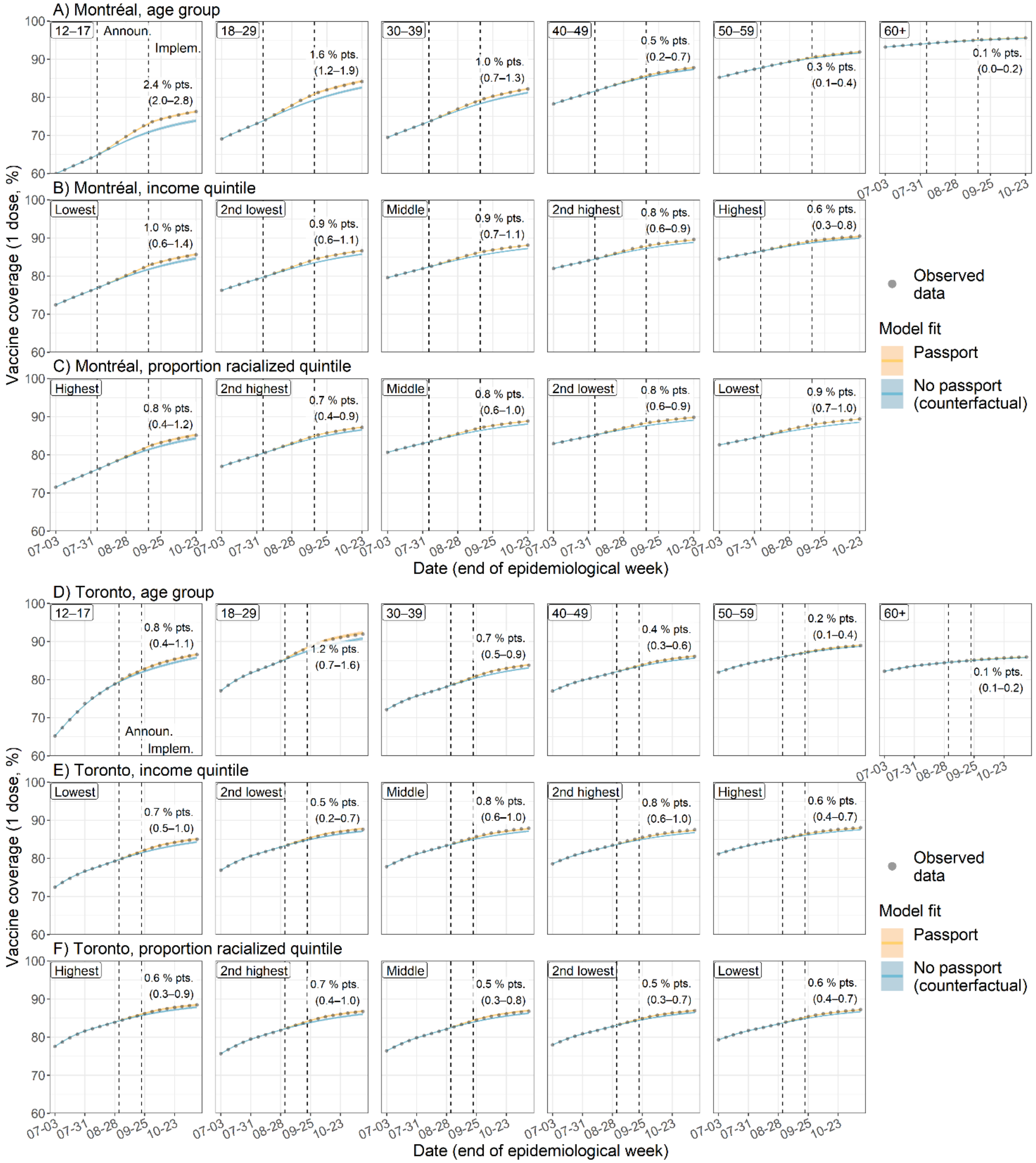


**Supplementary Figure S2. First-dose COVID-19 vaccine coverage in Montréal (A–C) and Toronto (D–F).** Observed (points) and modeled (blue and yellow) vaccine coverage over time is shown. Predicted vaccine coverage was obtained from three different regression models where the vaccination rate and the impact of the vaccine passport were allowed to vary by age group (A, D), DA-level income quintile (B, E), or DA-level proportion racialized quintile (C, F). Estimates and 95% confidence intervals (CIs) of the impact of the vaccine passport (observed coverage minus modeled counterfactual) are shown at the right of each panel. 95% CIs were estimated via bootstrap with 1,000 replicates. Announ., announcement of the vaccine passport; Implem., implementation of the vaccine passport.

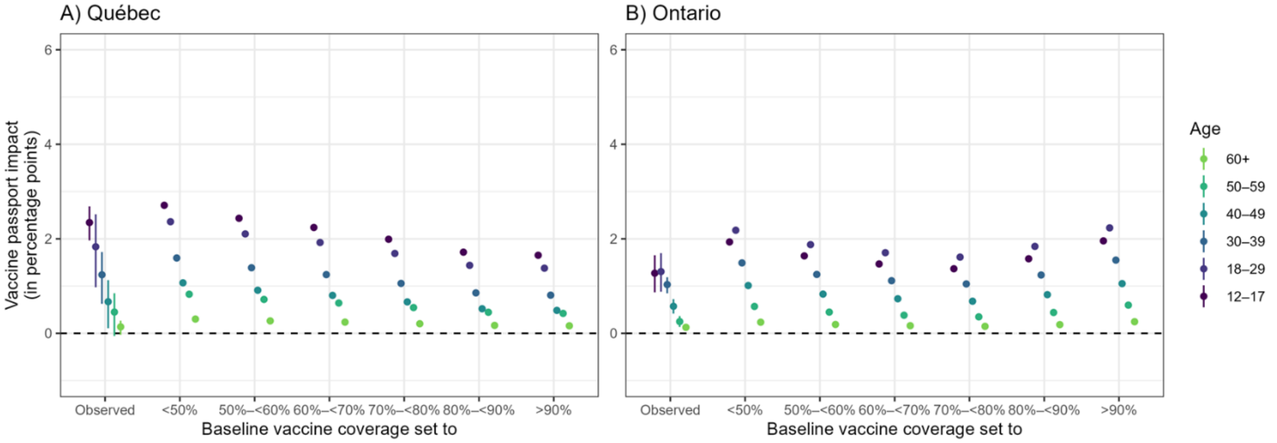


**Supplementary Figure S3. Impact of vaccine passport on first-dose coverage of COVID-19 vaccine (in percentage points) across age groups when holding baseline coverage constant for all dissemination areas (DA) in Québec (A) and Ontario (B) by the end of the study period.** The vaccine passport's impact (observed [or modeled] vaccination coverage minus the modeled counterfactual coverage in the absence of a vaccine passport) was estimated from a regression model where the vaccination rate and the impact of the vaccine passport were allowed to vary by age group and baseline coverage.

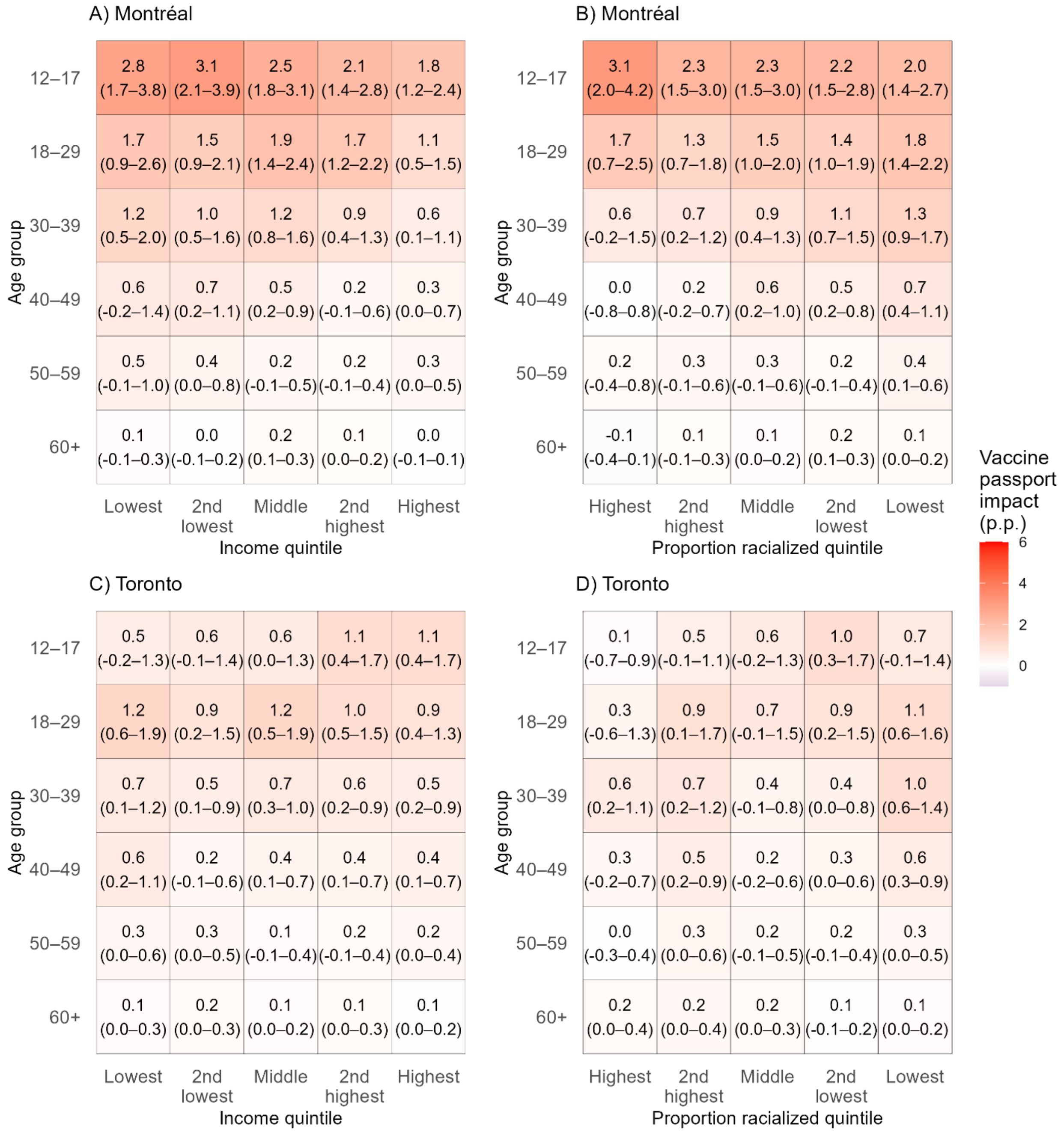


**Supplementary Figure S4. Impact of vaccine passport on first-dose coverage of COVID-19 vaccine (in percentage points) across age and by dissemination area (DA) level of income and proportion of racialized residents in Montréal (A, B) and Toronto (C, D) by the end of the study period.** The vaccine passport's impact (defined as the observed vaccination coverage minus the modeled counterfactual coverage in the absence of a vaccine passport) was estimated from two different regression models where the vaccination rate and the impact of the vaccine passport were allowed to vary by the interaction of age and either DA-level income quintile (A, C), or DA-level proportion racialized quintile (B, D). 95% confidence intervals –shown in parenthesis– were estimated via bootstrap with 1,000 replicates. p.p., percentage points.

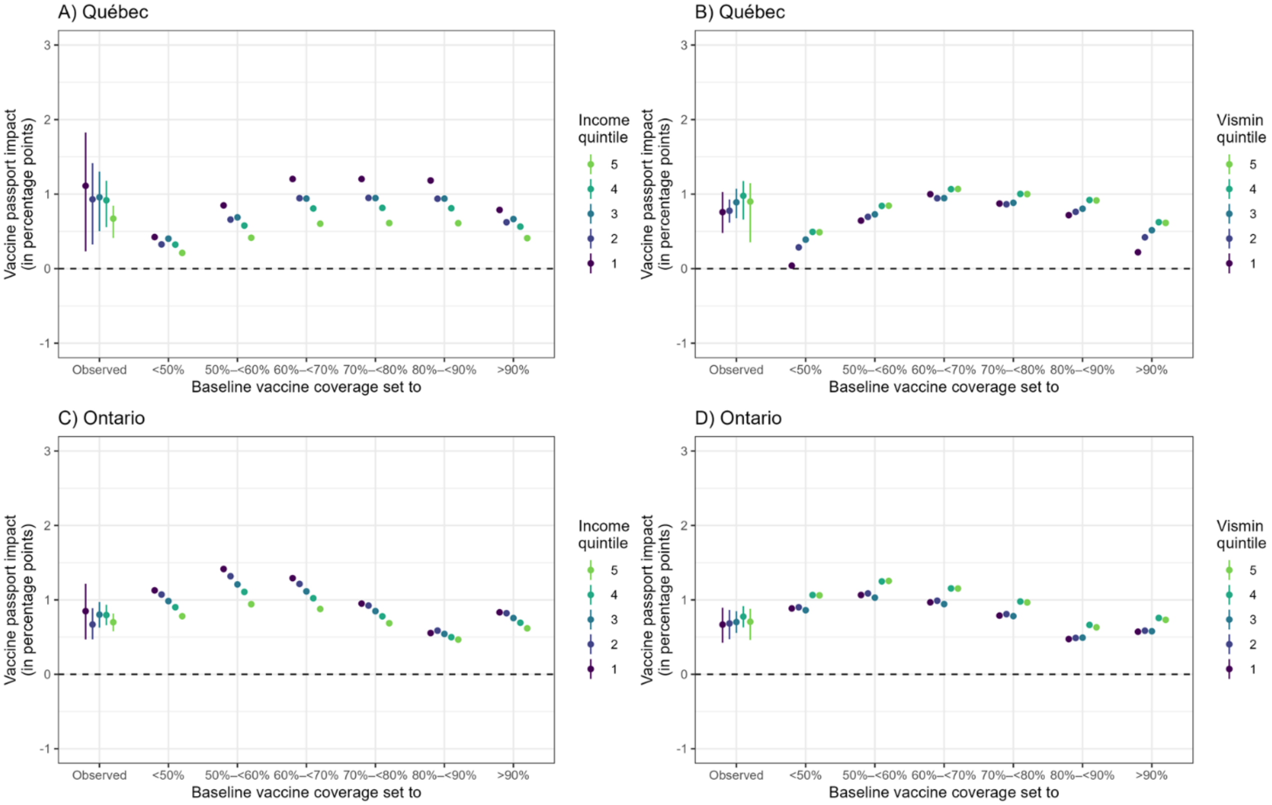


**Supplementary Figure S5. Impact of vaccine passport on first-dose coverage of COVID-19 vaccine (in percentage points) across social determinants when holding baseline coverage constant for all dissemination areas (DA) in Québec (A, B) and Ontario (C, D) by the end of the study period.** The vaccine passport's impact (observed [or modeled] vaccination coverage minus the modeled counterfactual coverage in the absence of a vaccine passport) was estimated from two different regression models where the vaccination rate and the impact of the vaccine passport were allowed to vary by baseline vaccine coverage and either DA-level income quintile (A, C), or DA-level proportion racialized quintile (B, D).

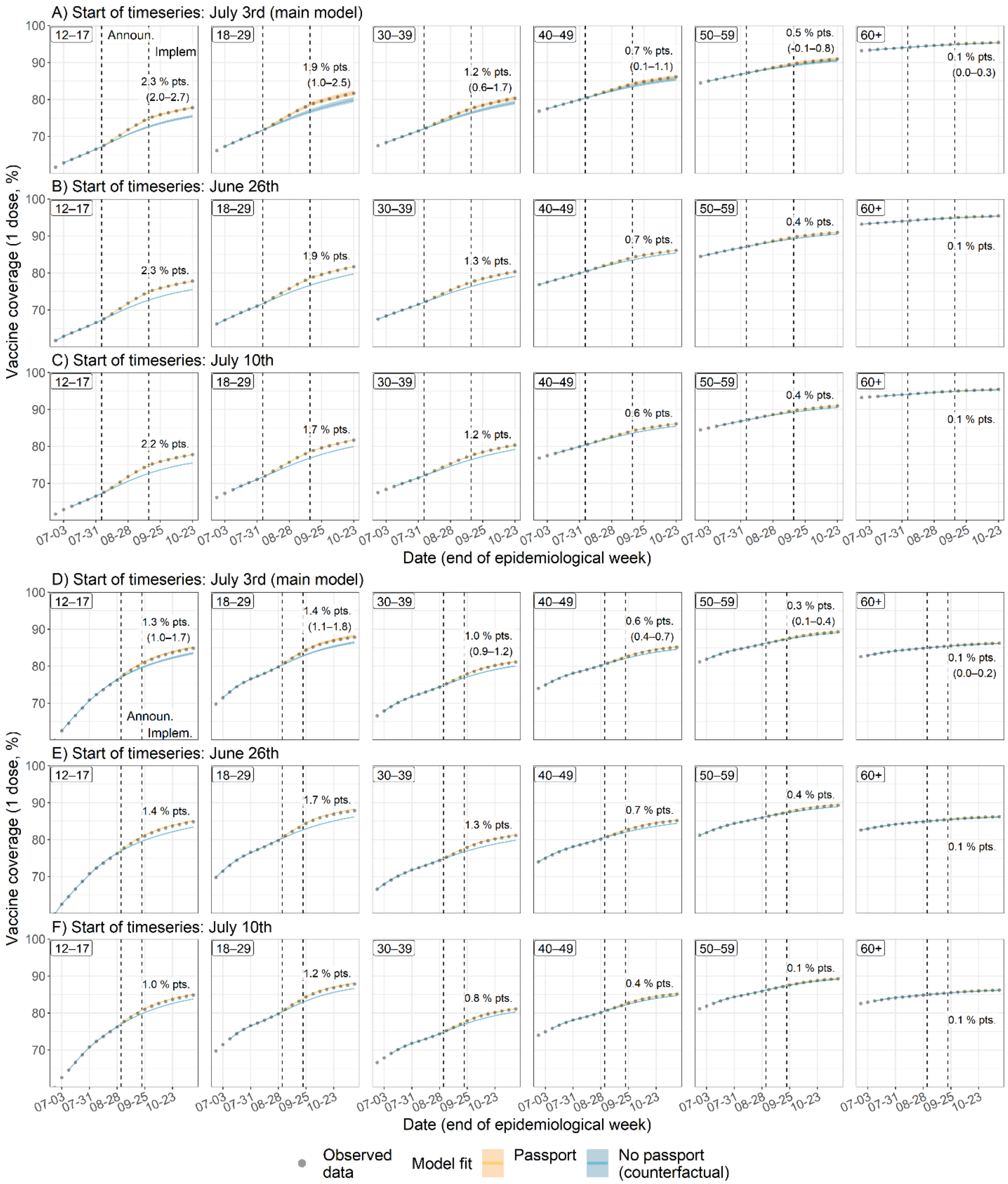


**Supplementary Figure S6. Impact of changing start of timeseries on first-dose COVID-19 vaccine coverage and the estimated vaccine passport effect in Québec (A–C) and Ontario (D–F).** Observed (points) and modeled (blue and yellow) vaccination rate over time is shown. Each row presents model fits from a different regression model, all of which allow the impact of the vaccine passport to vary by age group. Data for the regression models starts on July 3rd (main model; A,D), June 26th (B,E) or July 10th (C,F). Estimates and 95% confidence intervals (CIs) of the impact of the vaccine passport (observed coverage minus modeled counterfactual) are shown at the right of each panel. 95% CIs were estimated via bootstrap with 1,000 replicates. Announ., announcement of the vaccine passport; Implem., implementation of the vaccine passport.

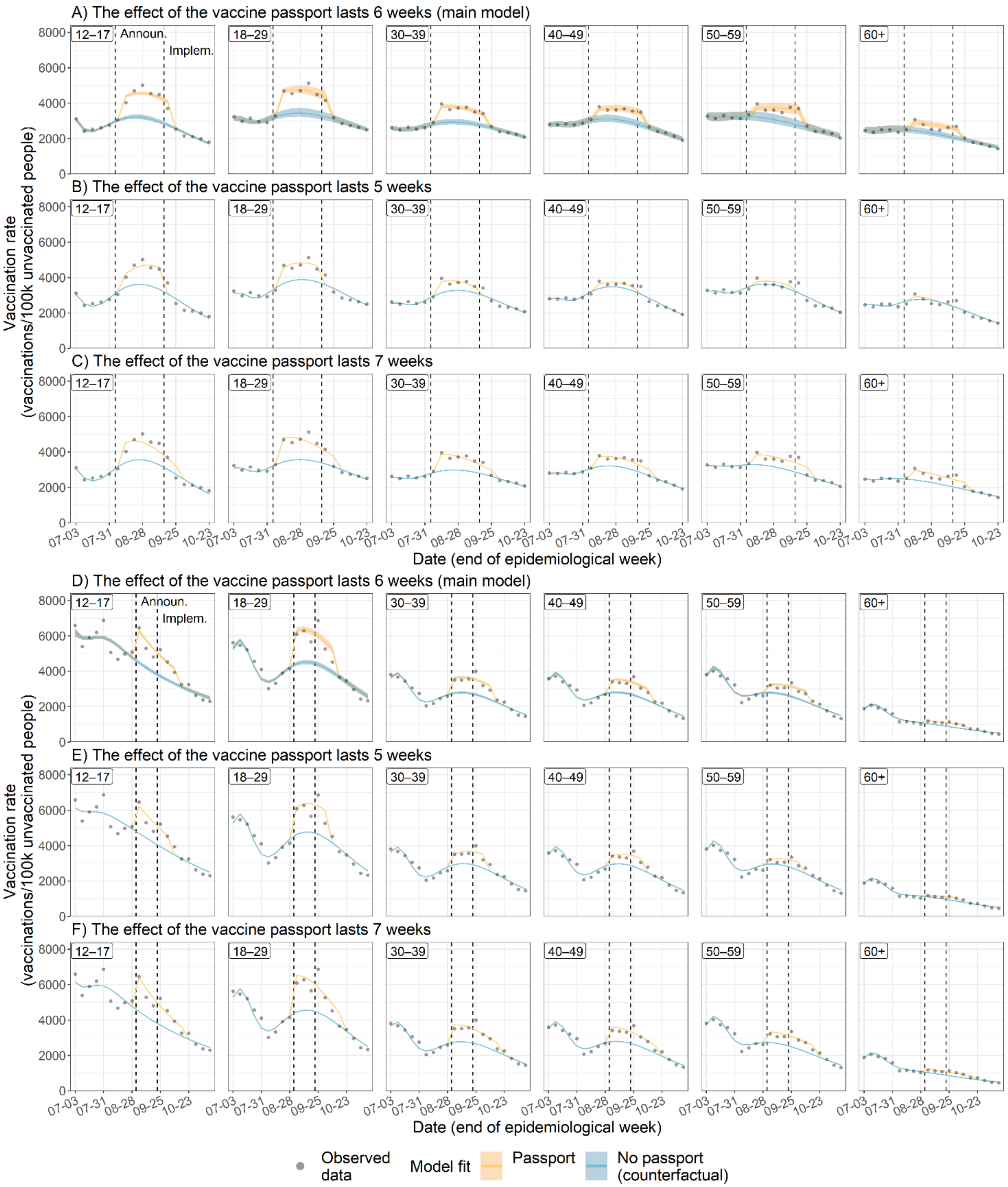


**Supplementary Figure S7. Impact of changing the length of the vaccine passport impact period on the weekly vaccination rate in Québec (A–C) and Ontario (D–F).** Observed (points) and modeled (blue and yellow) vaccination rates over time are shown. Each row presents model fits from a different regression model, all of which allow the impact of the vaccine passport to vary by age group. The vaccine passport was assumed to have an impact for a period of six weeks (main model; A,D), five weeks (B,E), or seven weeks (C,F). 95% confidence intervals were estimated via bootstrap with 1,000 replicates. Announ., announcement of the vaccine passport; Implem., implementation of the vaccine passport.

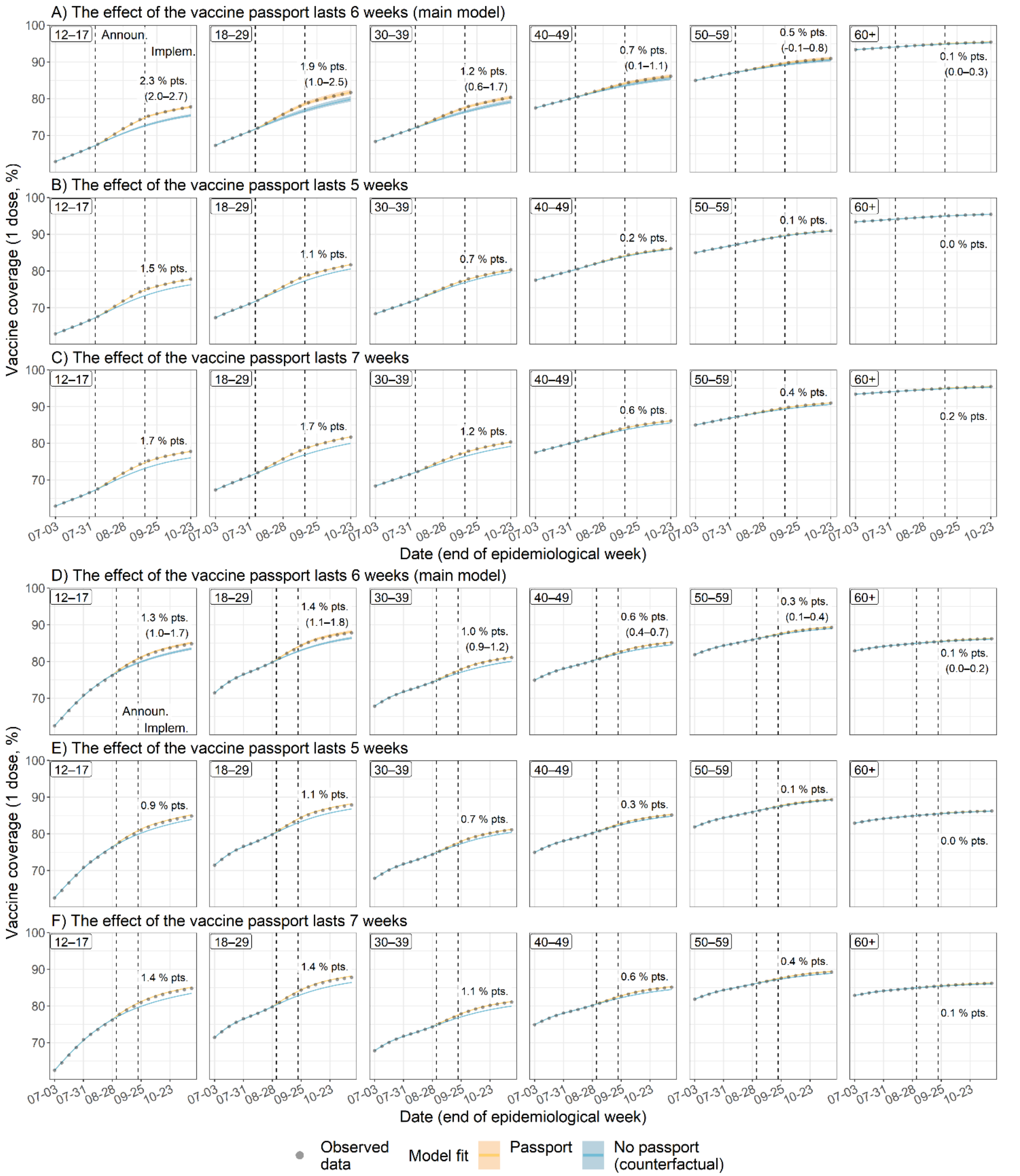


**Supplementary Figure S8. Impact of changing the length of the vaccine passport impact period on first-dose COVID-19 vaccine coverage and the estimated vaccine passport effect in Québec (A–C) and Ontario (D–F).** Observed (points) and modeled (blue and yellow) vaccination coverage over time is shown. Each row presents model fits from a different regression model, all of which allow the impact of the vaccine passport to vary by age group. The vaccine passport was assumed to have an impact for a period of six weeks (main model; A,D), five weeks (B,E), or seven weeks (C,F). Estimates and 95% confidence intervals (CIs) of the impact of the vaccine passport (observed coverage minus modeled counterfactual) are shown at top right of each panel. 95% CIs were estimated via bootstrap with 1,000 replicates. Announ., announcement of the vaccine passport; Implem., implementation of the vaccine passport.

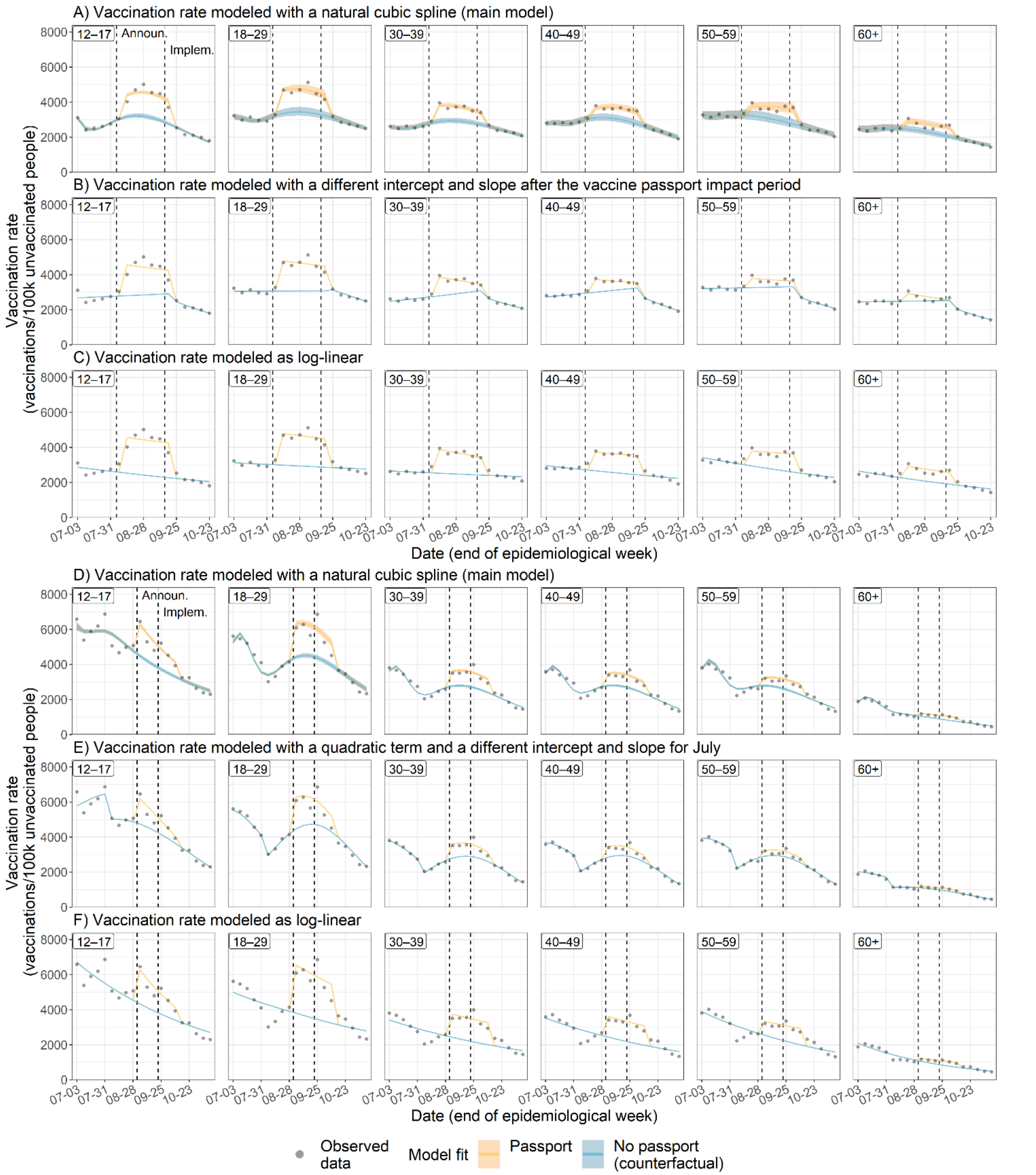


**Supplementary Figure S9. Impact of changing how the temporal trend is modeled on the weekly vaccination rate in Québec (A–C) and Ontario (D–F).** Observed (points) and modeled (blue and yellow) vaccination rates over time are shown. Each row presents model fits from a different regression model, all of which allow the impact of the vaccine passport to vary by age group. The vaccination rate-calendar time relationship is modeled with a natural spline (main model; A,D), a change in level and slope after the end of the vaccine passport's impact period (B), a quadratic term and a change in level and slope in July (D), or a log-linear relationship (C,F). 95% confidence intervals were estimated via bootstrap with 1,000 replicates. Announ., announcement of the vaccine passport; Implem., implementation of the vaccine passport.

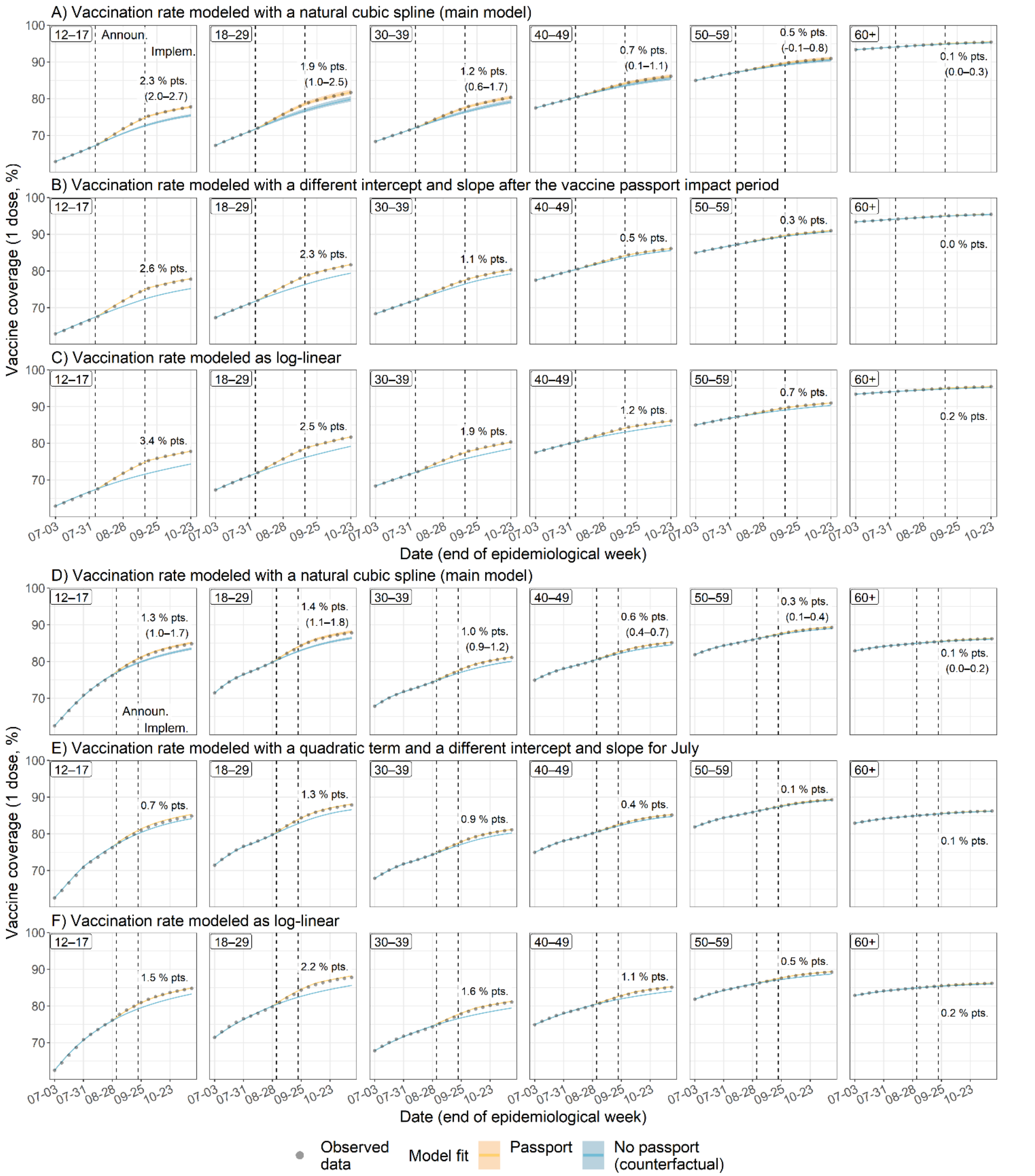


**Supplementary Figure S10. Impact of changing how the temporal trend is modeled on first-dose COVID-19 vaccine coverage and the estimated vaccine passport effect in Québec (A–C) and Ontario (D–F).** Observed (points) and modeled (blue and yellow) vaccination coverage over time is shown. Each row presents model fits from a different regression model, all of which allow the impact of the vaccine passport to vary by age group. The vaccination rate-calendar time relationship is modeled with a natural spline (main model; A,D), a change in level and slope after the end of the vaccine passport's impact period (B), a quadratic term and a change in level and slope in July (D), or a log-linear relationship (C,F). Estimates and 95% confidence intervals (CIs) of the impact of the vaccine passport (observed coverage minus modeled counterfactual) are shown at the right of each panel. 95% CIs were estimated via bootstrap with 1,000 replicates. Announ., announcement of the vaccine passport; Implem., implementation of the vaccine passport.

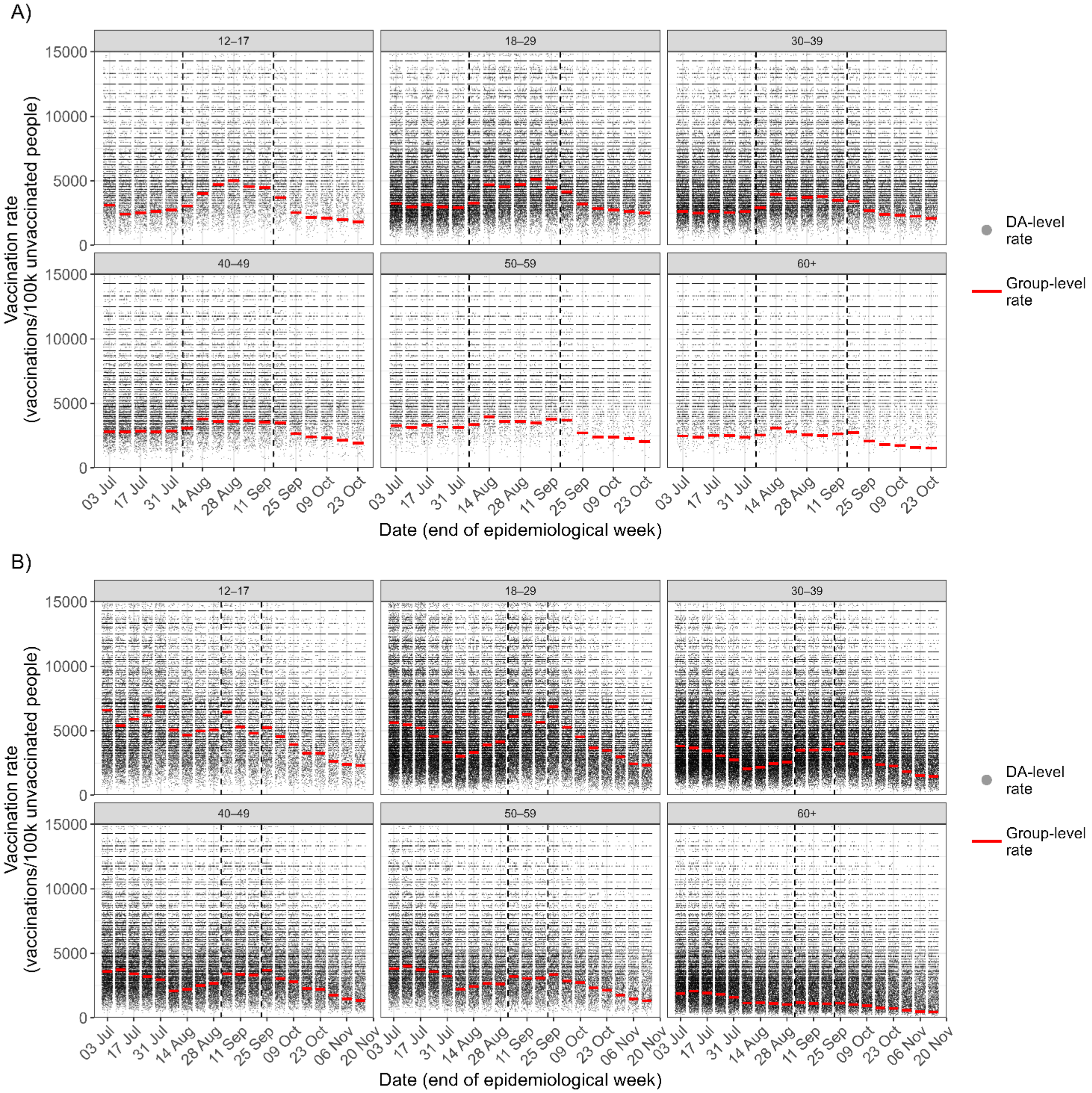


**Supplementary Figure S11. Distribution of the dissemination area (DA)-level weekly vaccination rate by age group in Québec (A) and Ontario (B).** Observed weekly vaccination rates over time are shown, each dot represents data from a single DA and each panel shows a different age group. DA, dissemination area.

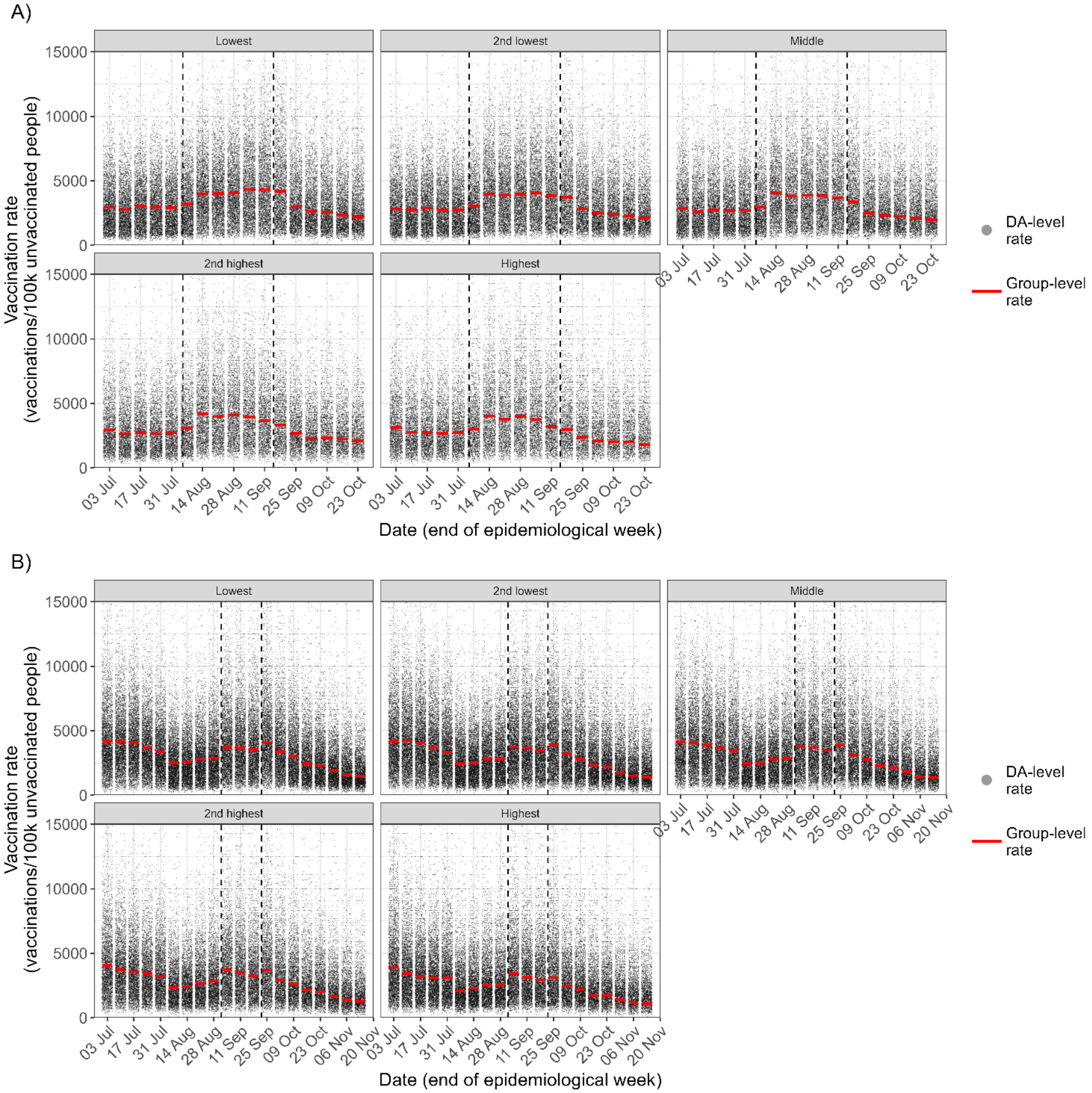


**Supplementary Figure S12. Distribution of the dissemination area (DA)-level weekly vaccination rate by income quintile in Québec (A) and Ontario (B).** Observed weekly vaccination rates over time are shown, each dot represents data from a single DA and each panel shows a different income quintile. DA, dissemination area.

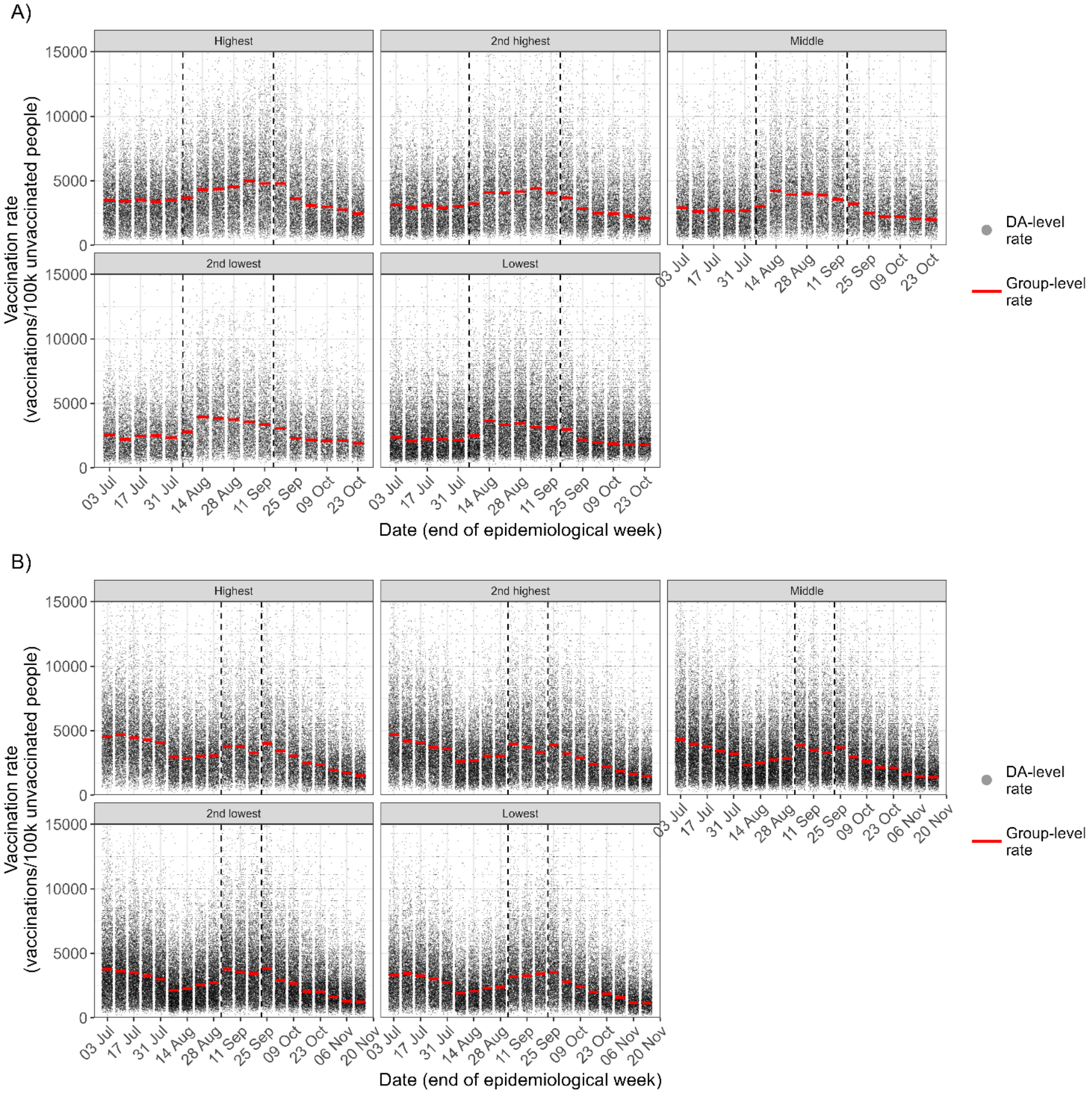


**Supplementary Figure S13. Distribution of the dissemination area (DA)-level weekly vaccination rate by proportion racialized quintile in Québec (A) and Ontario (B).** Observed weekly vaccination rates over time are shown, each dot represents data from a single DA and each panel shows a different quintile of proportion racialized. DA, dissemination area.
